# Hybrid adaptive immunity to SARS-CoV-2 protects against breakthrough infection after COVID-19 vaccination in ALSPAC participants

**DOI:** 10.1101/2024.06.14.24308948

**Authors:** Holly E. Baum, Marianna Santopaolo, Ore Francis, Emily Milowdowski, Katrina Entwistle, Elizabeth Oliver, Benjamin Hitchings, Divya Diamond, Amy C. Thomas, Ruth E. Mitchell, Milla Kibble, Kapil Gupta, Natalie Di Bartolo, Paul Klenerman, Anthony Brown, Begonia Morales-Aza, Jennifer Oliver, Imre Berger, Ash M. Toye, Adam Finn, Anu Goenka, Andrew D. Davidson, Susan Ring, Lynn Molloy, Melanie Lewcock, Kate Northstone, Firona Roth, Nicholas J. Timpson, Linda Wooldridge, Alice Halliday, Laura Rivino

## Abstract

Immunological memory to vaccination and viral infection involves coordinated action of B and T-cells, thus integrated analysis of these two components is critical for understanding their contributions to protection against breakthrough infections (BI). We investigated cellular and humoral immune responses to SARS-CoV-2 infection and/or COVID-19 vaccination in participants from the Avon Longitudinal Study of Parents and Children (ALSPAC). The magnitude of antibody and T-cell responses following the second vaccine dose was associated with protection against BI in participants with a history of SARS-CoV-2 infection (cases), but not in infection-naïve controls. Youden’s index thresholds for protection against BI were calculated for all immune measures. Anti-Spike IgG (>666.4 BAU/mL) and anti-Nucleocapsid pan Ig (>0.1332 BAU/mL) thresholds combined were 100% specific and 83.3% sensitive for cases without BI over 8-months follow-up. Collectively these results point to the superior protective effect of hybrid immunity and have implications for the design of next-generation COVID-19 vaccines.

## INTRODUCTION

The COVID-19 pandemic caused substantive medical and socioeconomic burdens that continue to affect the global population^1–4^. Infection with severe acute respiratory syndrome coronavirus 2 (SARS-CoV-2) presents as a broad spectrum of clinical manifestations from asymptomatic or mild infections, through to severe illness, and associated mortality^5,6^. The rapid deployment of COVID-19 vaccines proved instrumental in reducing SARS-CoV-2 infections and subsequent hospitalisations and deaths, however vaccine effectiveness wanes over time^7–9^. Breakthrough infections (BIs) are well documented and are influenced not only by waning immunity, but also the evolution of novel viral variants^10–13^.

A broad repertoire of antibody and cellular responses are elicited by SARS-CoV-2 infection^14–16^ and COVID-19 vaccination^17–19^. Evidence for anti-Spike (S) IgG and neutralizing antibody titres as correlates of protection (COPs) against SARS-CoV-2 infection and severe disease have been demonstrated in COVID-19 vaccine trials^20,21^, and studies of BIs^12,13^. Additionally, both systemic and mucosal IgA have been implicated in protection against infection^22–24^. SARS-CoV-2-specific T-cell responses are more durable than antibodies^25^, and may therefore be important for protection longer-term, and against novel variants which can evade antibody-mediated immunity^26,27^. Furthermore, cross-reactive T-cells can enhance infection- and vaccine-mediated responses^28^ and induce abortive infections in highly exposed individuals^29^.

Anti-viral immune memory responses are determined by the coordinated action of antibodies and T-cells targeting the virus, and an integrated analysis of these two components is therefore critical for understanding their contributions to protection. In this study we investigate the cellular and humoral responses to SARS-CoV-2 after infection and/or vaccination in participants from the Avon Longitudinal Study of Parents and Children (ALSPAC). We further address the role of antibodies and T-cells in providing protection against BI in participants with hybrid immunity, induced by a combination of SARS-CoV-2 infection and COVID-19 vaccination, as compared to those with vaccine-induced immunity alone.

## RESULTS

### Study design and recruitment

This study was open to ALSPAC participants of the G0 (48-70 years) and G1 (29-30 years) generations. 377 participants enrolled, and attended clinics in December 2020, March 2021, and June 2021 (Figure 1). Study participants were 59.2% female, 97.8% white, and 58.5% G1 generation (Table S1). Participants were selected from a cohort of 4819 individuals who had a valid SARS-CoV-2 antibody lateral flow test (LFT) result in October 2020. LFT anti-S IgG positivity rates were 3.1% and 5.8% in the G0 and G1 generations respectively (Figure 1). These results were used as an initial screen to identify participants with and without a likely history of SARS-CoV-2 infection for inclusion in this study^30^.

**Figure 1:**
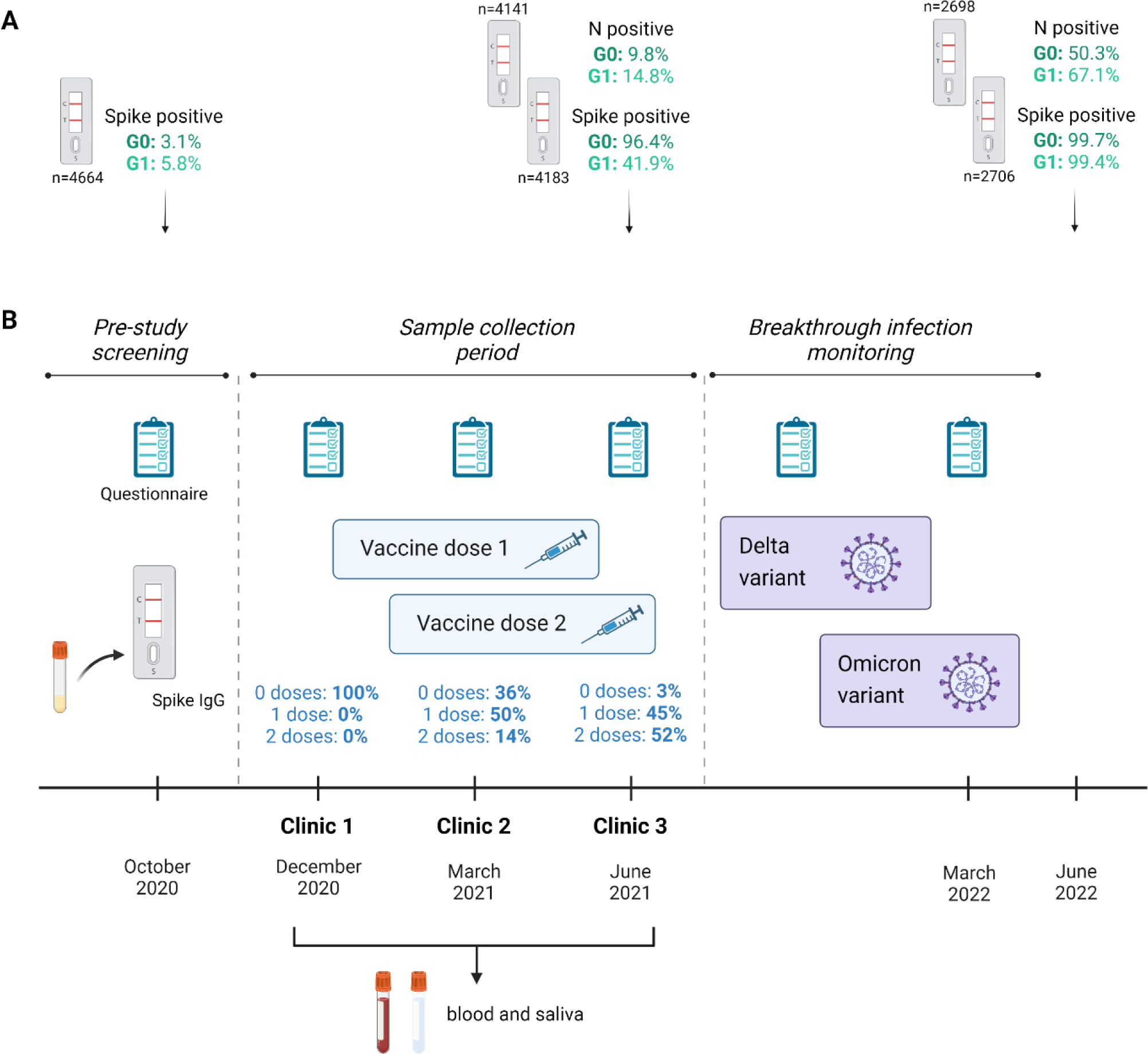
Study design. (A) Participants from the Avon Longitudinal Study of Parents and Children (ALSPAC) were screened for SARS-CoV-2-specific anti-Spike IgG in October 2020, prior to the vaccine rollout. Both the G0 (48-70 years) and G1 (29-30 years) generations were eligible for screening. Further sampling was conducted in June 2021 and June 2022 for both anti-Spike and anti-N IgG. Numbers indicate the total valid tests, with the percentages indicating the positivity rates. (B) Of those tested in October 2020, 377 were recruited to this study and attended one or more clinics from December 2020 to June 2021, providing biological samples, and completing health questionnaires. During this period, participants became eligible to receive COVID-19 vaccines via the UK national vaccination programme (blue text indicates % vaccinated at each clinic). Following the sampling period, participants continued to complete online questionnaires detailing their LFT and PCR-confirmed SARS-CoV-2 infections.

To validate the LFT results, serum samples from clinic 1 were screened for SARS-CoV-2-specific anti-S and anti-Nucleocapsid (N) antibodies using in-house ELISAs. Participants were defined as ‘cases’ if they had a previous PCR-confirmed SARS-CoV-2 infection and/or were positive on the anti-N or anti-Spike ELISAs (Figure 2A). The remaining seronegative individuals with no history of COVID-19 are herein referred to as ‘controls’. Clinic 1 samples were taken prior to participants being vaccinated as part of the UK COVID-19 vaccination programme. Of those who provided information on their vaccination status, 64.0% had received at least one dose by clinic 2, rising to 97.0% by clinic 3 (Figure 1). Data was therefore analysed relative to the number of COVID-19 vaccinations received (Figure S1), rather than chronologically by clinic, as this was deemed to be the dominant variable influencing immune responses.

**Figure 2:**
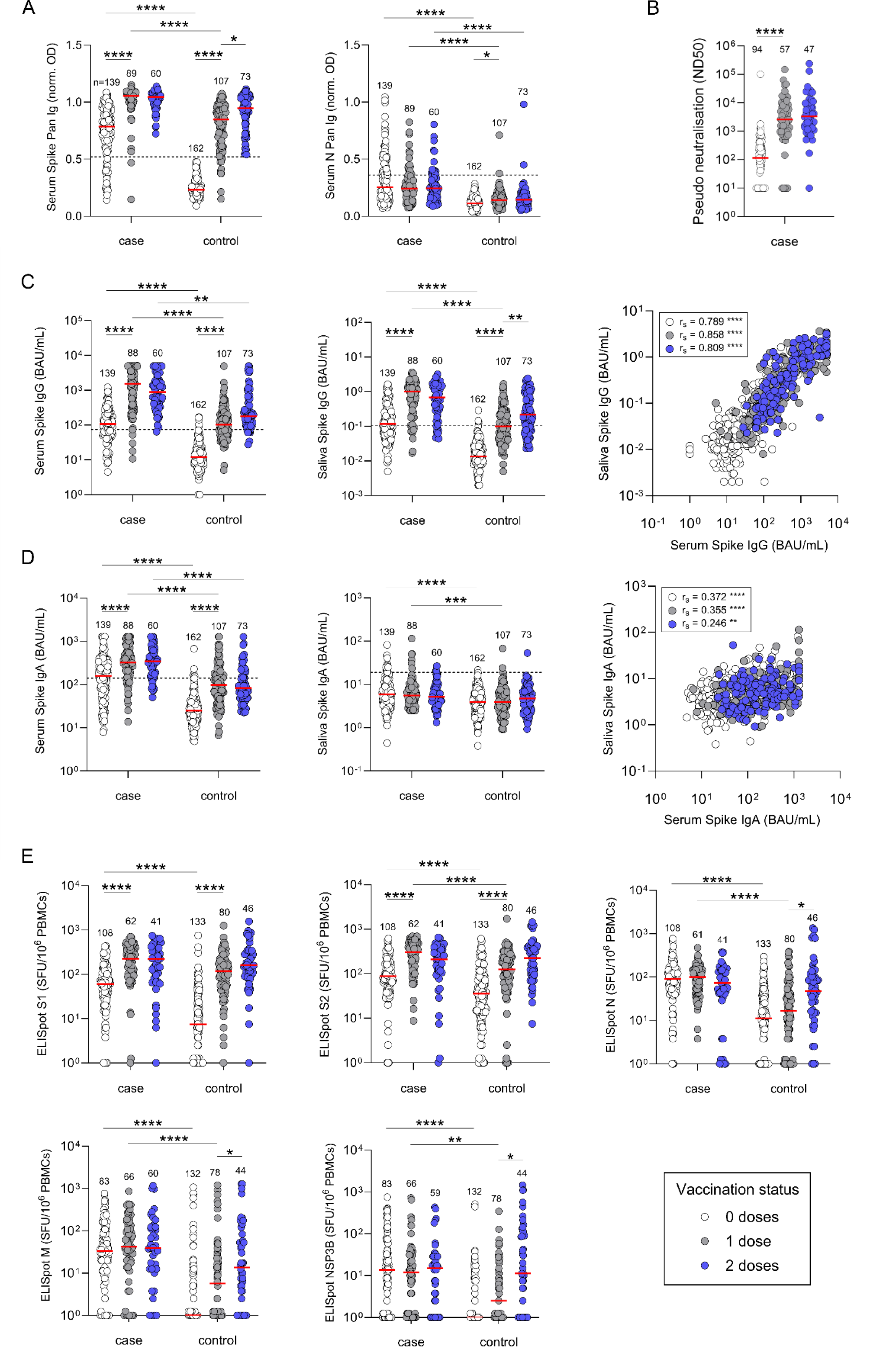
Antibody and T-cell responses to COVID-19 vaccination in SARS-CoV-2 naïve and previously infected individuals. (A) Participants were classified as cases based on a previous PCR-confirmed SARS-CoV-2 infection and/or positivity on serum anti-Spike and/or anti-Nucleocapsid pan Ig ELISAs. (B) Serum pseudoneutralising antibody titres. (C/D) Anti-Spike IgG and IgA in serum and saliva, measured by ELISA. Correlation coefficients calculated using Spearman’s rank (rs). (E) Magnitude of SARS-CoV-2-specific T-cell responses against Spike-1, Spike-2, N, M and NSP3B peptide pools measured by ELISpot assay. Red bars indicate median responses. Unpaired comparisons were performed using Kruskal-Wallis test with Dunn’s correction for multiple comparisons. Within each of the case and control groups, responses were compared between 0 and 1, and 1 and 2 vaccine doses. Responses between groups were compared after each dose. Statistics are only displayed for comparisons where P ≤0.05 (*), ≤0.01 (**), ≤0.001 (***), ≤0.0001 (****).

### Antibody responses to COVID-19 vaccination

At baseline, anti-S and anti-N pan Ig levels were higher in cases compared to controls (P≤0.0001; Figure 2A). A vaccine-induced increase in anti-S pan Ig was observed in both groups. Following the second dose, all participants had anti-S pan Ig levels above the assay positivity threshold. However, the case group retained their baseline advantage with higher median titres. Rates of new SARS-CoV-2 infections between clinics 1-3 were low, with four individuals in the control group reporting a positive PCR test. Consistent with this observation, a small increase in median anti-N pan Ig was observed after the first dose (P=0.0471), which plateaued after dose 2 (Figure 2A). This was predominantly driven by ≥2-fold increases in anti-N pan Ig levels in a small minority of participants (3/95; Figure S2). Anti-N pan Ig levels in the case group remained stable throughout.

Anti-S IgG in serum and saliva showed similar kinetics to serum anti-S pan Ig responses. Post-vaccination anti-S IgG levels in the two sample types were very strongly correlated after the first (r_s_=0.858, P≤0.0001) and second (r_s_=0.809, P≤0.0001) doses (Figure 2C). Serum pseudoneutralising antibody titres in the case group increased in response to the first vaccination, then remained stable following the second dose (Figure 2B). A strong positive correlation between anti-S IgG and pseudoneutralising antibody titre was observed following each dose (r_s_=0.784, P≤0.0001 / r_s_=0.656, P≤0.0001; Figure S3).

Pre-vaccination anti-S IgA in serum and saliva was higher in cases than controls (Figure 2D). Serum anti-S IgA increased in both groups after the first dose, whilst median salivary anti-S IgA levels were not boosted by vaccination. There was therefore only a weak correlation between the two measures post-vaccination (r_s_=0.355, P≤0.0001 / r_s_=0.246, P=0.0045). Recipients of mRNA vaccines had higher median serum anti-S IgG and IgA levels than recipients of the adenoviral vector based ChAdOx1 (Figure S4).

### T-cell responses to COVID-19 vaccination

T-cell responses targeting SARS-CoV-2 S1/S2, N, Membrane (M), Envelope (E), non-structural proteins (NSP3-16) and accessory proteins (ORF3, ORF6, ORF7 and ORF8) were measured by IFN-γ ELISpot (Figure S5A/B). Baseline S-specific IFN-γ responses were lower in controls compared to cases (P≤0.0001; Figure 2E). In both groups, S-specific T-cells increased after the first vaccination and then remained stable. In the case group, T-cell responses to N, M and NSP3B were not boosted by vaccination while those to NSP 1+2 increased after vaccination. In contrast, increased T-cell responses to M (P≤0.0001), N (P=0.0175), and NSP3B (P=0.0091) were observed in control participants after dose 2 compared to baseline (Figure 2E). T-cell responses to NSP3-16, ORF3, ORF7 and ORF8, remained stable following vaccination (Figure S5A/B).

### Magnitude and quality of T-cell responses to SARS-CoV-2 infection

The magnitude and functional phenotype of the CD4^+^ and CD8^+^ T-cell response targeting S, N, M and NSP3B proteins were analysed in cases, prior to vaccination. Production of IFN-γ, TNF-α, MIP1β, IL-2 and degranulation (CD107a) by SARS-CoV-2-specific CD4^+^ and CD8^+^ T-cells was measured by ICS. A representative gating strategy is shown in Figure S6. Responses were comprised of monofunctional and polyfunctional T-cells producing 1 or >1 effector functions respectively, with monofunctional T-cells dominating both the CD4^+^ and CD8^+^ T-cell response (Figure 3; see Table S2 for statistical comparisons). Monofunctional SARS-CoV-2-specific CD4^+^ T-cells produced mainly TNF-α (S1=59.03%, S2=28.45%, M=25.7%, N=41.35%, NSP3B=27.35%) and IFN-γ (S1=23.59%, S2=16.75%, M=35.39%, N=34.55%, NSP3B=48.34%). The frequencies of CD4^+^ T-cells producing each cytokine were comparable across proteins (Figure S7A). In contrast, CD107a was expressed at higher levels by CD4^+^ T-cells targeting M (34.26%), compared to S1 (5.25%), S2 (5.23%), and N proteins (9.42%). Polyfunctional CD4^+^ T-cells were mainly IFN-γ^+^/TNF-α^+^, IL-2^+^/TNF-α^+^ and IFN-γ^+^/IL-2^+^/TNF-α^+^ (Figure 3A).

**Figure 3:**
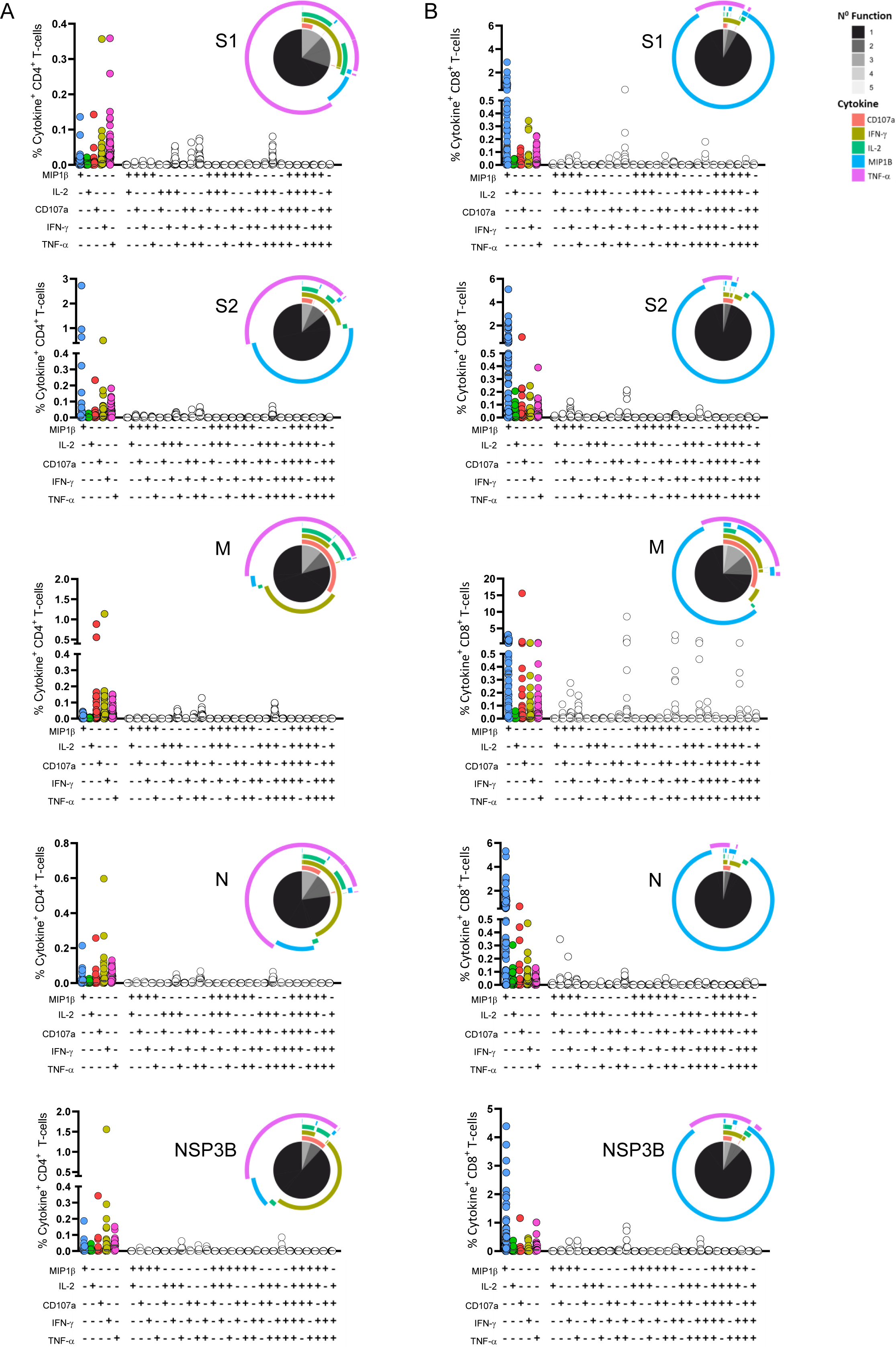
SARS-CoV-2-specific CD4^+^ and CD8 ^+^ T-cell responses in previously infected individuals. Graphs show the percentage of CD4^+^ (A) and CD8^+^ (B) T-cells producing the indicated cytokine or combination of cytokines after a brief stimulation with Spike-1, Spike-2, M, N or NSP3B peptide pools, assessed by intracellular cytokine staining and flow cytometry. Pie charts indicate the proportion of T-cells, within total cytokine^+^ T-cells, that producing each cytokine (colour-coded) and that display one or more functions (black/grey). Statistics from monofunctional SARS-CoV-2-specific CD4^+^ and CD8^+^ T-cells in previously infected individuals were calculated using a Kruskal-Wallis test with FDR method of Benjamini and Hochberg correction for multiple comparisons. P-values are reported in Table S2.

Similarly to CD4^+^ T-cells, cytokine production by CD8^+^ T-cells targeting the different SARS-CoV-2 proteins was comparable while CD107a was expressed by a larger proportion of CD8^+^ T-cells specific for M compared to other proteins (M=31.7%, S1=1.63%, S2=4.91%, N=3.52%, NSP3B=4.27%) (Figure S7B). Polyfunctional CD8^+^ T-cells were mainly double or triple-cytokine producing, including IFN-γ^+^/TNF-α^+^, CD107a^+^/TNF-α^+^, MIP1β^+^/TNF-α^+^, MIP1β^+^/IFN-γ^+^ and MIP1β^+^/IFN-γ^+^/TNF-α^+^. NSP3B-specific CD8^+^ T-cells displayed higher polyfunctionality (14.8%) compared to those targeting S (S1=10.4%, S2=5.29%) or (N=5.33%) (Figure 3B; Figure S8A/B).

### Association between post-vaccination immune responses and protection against SARS-CoV-2 BI

We next investigated the relationship between the magnitude of antibody and T-cell responses after the second vaccination, and protection against SARS-CoV-2 BI. The case and control groups were subdivided based on self-reported questionnaire data detailing whether the participant had a PCR/LFT-confirmed SARS-CoV-2 infection in the 8-months following the sample collection period (July 2021 - March 2022; Figure 1). For this analysis, the case group was updated to include two individuals who had PCR-confirmed SARS-CoV-2 infections between clinics 1-3. BI rates were higher in the control group (42.4%) compared to the cases (30.8%). Of the individuals who reported BIs, the median time since receiving a second vaccination, and the proportion who received a third vaccination prior to infection, were comparable between cases and controls (Table S3). Self-reported BIs were consistent with increased rates of LFT anti-N IgG positivity detected in the wider ALSPAC cohort in the period from May 2021 (G1=14.8%; G0=9.8%) to May 2022 (G1=67.1%; G0=50.3%; Figure 1).

In participants without a history of COVID-19, there was no strong evidence of differences between those who went on to experience a BI, compared to those who didn’t, with respect to the magnitude of immune variables measured (Figure S9A; Figure 4A). Conversely, previously infected participants showed evidence of clustering by subsequent infection status (Figure 4B). Higher levels of anti-S IgG in serum and saliva, pseudoneutralising antibody, and anti-S IgA in serum as well as S1-specific T-cell responses were observed in those with no reported BI (Figure S9B). Similarly, high anti-N Pan Ig titres were associated with a decreased likelihood of re-infection. The lack of vaccine-induced salivary IgA in this cohort was reflected in the poor performance of this measure as a discriminator of future infection susceptibility.

**Figure 4:**
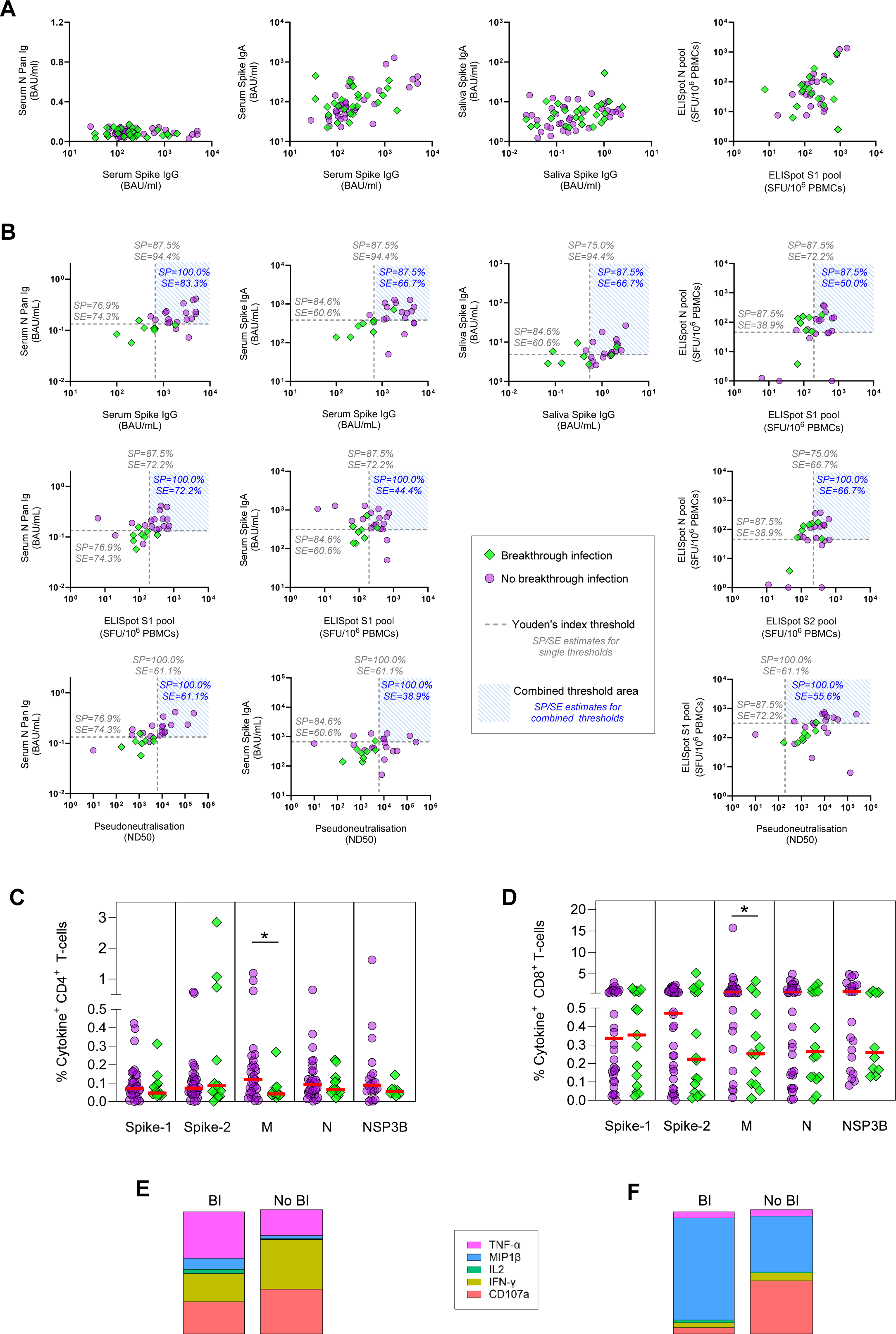
Association between immune responses to prior SARS-CoV-2 infection and COVID-19 vaccination and susceptibility to SARS-CoV-2 breakthrough infection. (A/B) Correlation of post-second vaccination antibody and T-cell responses in participants who self-reported a SARS-CoV-2 infection in the subsequent 8-months (breakthrough infection; BI) and those who didn’t (no BI). (A) SARS-CoV-2-naïve individuals (with BI n=14; no BI n=19). (B) Previously SARS-CoV-2-infected individuals (BI n=8, no BI n=18). See Figure S10 for derivation of thresholds. (C-F) Correlation of pre-vaccination SARS-CoV-2-specific T-cell intracellular cytokine responses in cases with subsequent BI. (C/D) Magnitude of baseline (pre-vaccination) single cytokine/CD107a-producing (monofunctional) SARS-CoV-2-specific CD4^+^ (C; n=47) and CD8^+^ (D; n=51) T-cells specific for the indicated SARS-CoV-2 proteins among cases. Red bars represent median responses. Statistics calculated by t-test (Mann-Whitney). (E/F) Proportions of single cytokine/CD107a-producing CD4^+^ (n=40) and CD8^+^ (n=43) T-cells within the respective M-specific T-cell population.

To compare the effectiveness of individual markers for discriminating between cases who did or didn’t go on to be re-infected, ROC curves were plotted, and thresholds calculated using the Youden’s Index method to balance sensitivity and specificity (Figure S10). Anti-S IgG ≥666.4 BAU/mL in serum (SP=87.5%), or 0.547 BAU/mL in saliva (SP=75.0%), provided 94.4% sensitivity for identifying participants who didn’t report BIs (Figure 4). Specificity was improved to 100.0% by combining serum or saliva S-specific IgG, and N-specific pan Ig, thresholds (SP=100%, SE=83.3%). Combining S-specific serum IgG with IgA in serum or saliva, or with T-cell responses, did not improve discrimination. Pseudoneutralising antibody ≥6165 units provided a highly specific threshold (SP=100%) for identification of protected individuals, albeit with lower sensitivity (SE=61.1%) than the S-specific IgG threshold (SE=94.4%).

The magnitude of T-cell responses to the S1 pool (SP=87.5%, SE=72.2%) provided a more specific and sensitive marker of infection susceptibility than those to the S2 (SP=75.0%, SE=66.67%) or N (SP 87.5%, SE=38.9%). Combining thresholds for T-cell S1 and N did not improve performance compared to S1 alone. Conversely, combining T-cell S1 with N-specific pan Ig offered improved specificity with no loss of sensitivity (SP=100%, SE=72.2%). The performances of all threshold combinations are detailed in Figure S11.

### Association between ICS responses to pre-vaccination SARS-CoV-2 infection and BI

To determine whether specific features of the T-cell response to SARS-CoV-2 infections were also a factor in determining risk of BI, pre-vaccination ICS responses were compared in cases with and without a reported BI. Monofunctional CD4^+^ and CD8^+^ T cell responses were included in these analyses as these were the predominant responses observed in these individuals. There was no meaningful difference between the magnitude of S, N or NS3PB-specific T-cell responses in the two groups (Figure 4C/D, Figure S8C/D). In contrast, participants who didn’t report BIs displayed higher magnitudes of SARS-CoV-2 M-specific CD4^+^ and CD8^+^ T-cells, and these were skewed respectively towards production of IFN-γ^+^ and CD107a (IFN-γ^+^: 40.0%; CD107a: 35.7%; Figure 4E), and CD107a and MIP1β (CD107a: 42.4%; MIP1β: 45.0%; Figure 4F), respectively. However, including these ICS measures in the analysis did not improve the predictive ability of thresholds generated using antibody and/or T-cell data alone.

## DISCUSSION

Humoral and cellular immune responses to COVID-19 vaccination were measured in ALSPAC participants with or without a history or serological evidence of SARS-CoV-2 infection. In those with hybrid immunity, we demonstrate a correlation between the magnitude of responses following the second vaccination and protection against BI. The combination of serum S-specific IgG (>666.4 BAU/mL) and N-specific pan Ig (>0.1332 BAU/mL) thresholds identified those who didn’t report BIs with 100% specificity and 83% sensitivity. Our results suggest that hybrid immunity to SARS-CoV-2 remains effective in protection from reinfection at >15 months post infection and >8 months post second-dose vaccination. The reduced association in control participants alludes to the importance of the greater quality and breath of immune responses elicited by SARS-CoV-2 infection compared to vaccination alone.

COVID-19 vaccination elicited robust serum anti-S IgG and IgA production in all participants, with the case group retaining their baseline immunological advantage as observed in other studies^31^. mRNA vaccination resulted in higher S-specific IgG and IgA compared to the viral vector ChAdOx1, as previously reported^32^. However, neither vaccine type induced salivary IgA, even in participants with a history of COVID-19 where vaccination has been suggested to boost infection-primed responses^22,33,34^. Reducing or eliminating transmission with vaccines capable of eliciting mucosal immunity therefore remains a priority, and a number of candidates are currently in development^35^.

Consistent with recent papers, COVID-19 vaccines induced S-specific T-cell responses in all participants^36–38^. An increase in the median frequency of T-cell responses to M, N, and NSP3B was observed in the control group. This was predominantly driven by large increases to all target antigens in a small number of participants, suggestive of SARS-CoV-2 infections in these individuals during the study period. No parallel increases in serum anti-N pan Ig or saliva anti-S IgA were observed. These participants with may therefore have experienced asymptomatic, abortive SARS-CoV-2 infections which have been shown to boost T-cell responses in the absence of seroconversion^29^. Alternatively, these data could potentially suggest bystander T-cell activation as a result of vaccination. Antigen-independent bystander activation of T-cells which is likely driven by cytokines, has been widely reported during viral infection^39,40^, however its occurrence in the context of vaccination remains unclear^41–45^.

Long-term protection against SARS-CoV-2 BI relies upon the durability of vaccine-induced responses. Antibodies in particular have been shown to wane rapidly in the 6-months following the primary and subsequent vaccine doses^46–48^. This declining immunity, particularly in the context of highly transmissible SARS-CoV-2 variants with reduced neutralising sensitivity, increases BI rates^8,10,49^. Anti-S IgG and neutralising antibody levels have been shown to negatively correlate with the risk of SARS-CoV-2 infection and severe disease^12,21,50–52^. In control participants, we observed no evidence of difference in the magnitude of responses between those who did and didn’t experience BIs, and protective thresholds could therefore not be defined. However, anti-S IgG ≥666.4 BAU/mL was associated with reduced incidence of BI in those with a history of SARS-CoV-2 infection. This is consistent with the large Israeli COVID-19 family study (ICoFS) which proposed S-specific IgG >500 BAU/mL, and neutralising antibody titres of >1024, as thresholds of protection against SARS-CoV-2 Delta infection ^53^. IgG levels above this were associated with an 11% probability of infection, and 1% probability of moderate disease. ICoFS baseline measurements were taken shortly before infection and therefore their lower threshold value is expected relative to our use of post-vaccination dose 2 titres that were recorded months earlier. Serum anti-S IgA also associated with reduced BIs in our cohort, and alongside mucosal IgA has previously been shown to correlate with protection against infection independently of IgG^22^. BIs experienced shortly after primary vaccination correlated with lower vaccine-induced IgA^22^, and in those who experienced infections serum IgA levels inversely correlated with symptom duration^54^. Increased S-specific T-cells also correlated with reduced BI in our cohort, consistent with previous reports for both CD4^+^ and CD8^+^ responses^55,56^. Furthermore, the combination of high neutralizing antibodies and S-specific IFN-γ responses is associated with protection against BI^57,58^.

There is now mounting evidence that hybrid immunity affords more robust and sustained protection against BI compared to vaccine-induced immunity alone^46,59–62^. Consistent with this, BI rates in our study were lower in cases (30.8%) compared to controls (42.4%). Both humoral and cellular immune responses have been reported to be qualitatively superior in those who experience SARS-CoV-2 infection prior to vaccination^61,62^, and infection-derived immunity may be particularly important in the context of BIs with novel variants^23,63^. It is only SARS-CoV-2 infection that induces a significant and durable systemic IgA response^48^, and COVID-19 vaccines perform poorly at eliciting mucosal IgA^34^. Additionally, the increased magnitude of neutralising responses in hybrid immunity enhances the breadth of neutralising activity against divergent variants of concern^64,65^. Vaccine-induced Fc-receptor binding antibodies are also important for the control and clearance of infection and have also been shown to be more abundant in those with a history of COVID-19, with this difference persisting post-vaccination^61^.

Functional and phenotypic properties of SARS-CoV-2 adaptive immunity appear to differ in response to SARS-CoV-2 infection and COVID-19 vaccination^66,67^. A skewed T-helper (Th)1 SARS-CoV-2-specific T-cell response and higher percentages of IgG-expressing memory B-cells were observed in vaccinated individuals compared to individuals who recovered from COVID-19^68^. In this study we assessed the functional and phenotypic features of SARS-CoV-2-specific CD4^+^ and CD8^+^ T-cells in previously infected individuals prior to vaccination, and compared these in individuals who subsequently did or did not experience BIs. We show trends towards higher magnitudes of SARS-CoV2-specific CD8^+^ T-cells targeting S2, N, M and NSP3B in individuals who do not develop BI, with differences being significantly higher only for M, suggesting a protective role for these cells. Similarly, M-specific CD4^+^ T-cells are present at higher magnitudes in individuals who do not develop BI compared to those that do. In addition, the former group displays M-specific CD4^+^ T-cells mainly producing IFN-γ and CD107a, which suggests a protective role of cytotoxic Th1-cells in COVID-19. These data highlight how a granular analysis of CD4^+^/CD8^+^ T-cell responses may inform T-cell features associated with protection in COVID-19. This is critical for an integrated definition (incorporating T-cells and antibodies) of correlates/determinants of protective immunity to SARS-CoV-2 and potentially other viral infections.

In addition to changing the quality of responses, SARS-CoV-2 infection primes the immune system with a broader range of antigens than vaccination. Anti-N seropositivity was strongly associated with protection against reported BIs in our cohort. This corroborates the conclusions of other studies, with previous estimates suggesting that it afforded participants with approximately 80% protection against SARS-CoV-2 re-infection for the subsequent 8-months ^69^. In those experiencing BIs, anti-N seropositivity also associated with a shorter duration and reduced viral load^54^. Evidence suggests that COVID-19 vaccination drives differential responses to subsequent SARS-CoV-2 infection, boosting those primed by the vaccine over new responses to the broader repertoire of proteins in the virus^54,70–72^. Seroconversion to N upon SARS-CoV-2 infection was observed at a higher rate in placebo vaccinated participants (93%) compared to mRNA-1273 recipients (40%)^70^. Additionally, SARS-CoV-2 BIs elicit anti-N IgG at significantly lower rates in infection-naïve individuals compared to those with a history of pre-vaccination COVID-19^54,71,72^. This suggests that the order in which hybrid immunity is achieved may be important.

SARS-CoV-2 LFT testing of a wider sample of ALSPAC participants highlighted an age-related difference in anti-N positivity rates. By spring 2022, all UK adults had been offered 3 vaccine doses, and this was reflected in comparable anti-S positivity rates of 93.0% and 96.0% in the G0/G1 cohorts respectively. However, only 50.3% of the older generation had a positive anti-N response compared to 67.1% of the younger adults. These values may underestimate SARS-CoV-2 infection rates in the older generation due to the lower N-seroconversion rates in infections occurring post-vaccination. However, they do accurately reflect a lack of potentially protective anti-N antibodies in this population. Prioritising older adults for early vaccination, and encouraging shielding behaviours, may therefore have consequences for the breadth and durability of the immunity they obtain by reducing pre-vaccination infection rates, and overall N-seroconversion. Rapid vaccination of this age group proved essential for reducing the rates of severe disease and associated mortality, and on a population level these benefits are unlikely to be outweighed by the benefits of a broader infection-primed response. Instead, this highlights the importance of designing novel vaccines that prime responses to additional antigens to generate more robust and sustained protection.

Our BI susceptibility analysis should be interpreted in the context of the limitations of our study design. Firstly, the sample size was limited by those with available post-dose 2 vaccination measures and may not be representative of the whole population. Secondly, with respect to the BI rates in the case and controls groups, differences in exposure risk and test-seeking behaviours which could not be measured or accounted for may have influenced outcomes. Key variables, such as the proportion who received a booster vaccine, were however comparable between groups. Thirdly, N-protein serology does not offer 100% sensitivity for detection of new SARS-CoV-2 cases and therefore undetected infections during the study period could have resulted in participants being mis-classified as controls for the BI analysis. Fourthly, samples were not collected at the point of BI and therefore could not be confirmed virologically, nor the SARS-CoV-2 sequences/variant lineages determined. The BI infection monitoring period covered the boundary of Delta and Omicron being the predominant variants in circulation. For the purposes of this analysis, all SARS-CoV-2 infections have been analysed together, but it is possible that variant-specific thresholds for protection may differ. Fifthly, although we report correlations between immune markers and susceptibility to future infection, we cannot exclude the possibility that rather than being directly protective, some may act as proxies for undetermined variables that weren’t measured. Finally, sample size limitations prevented the stratification of the BI analysis by vaccine type. However, this has previously been shown not to directly influence the susceptibility to BIs. Instead, higher BI rates in ChAdOx1 recipients can be attributed to the lower immunogenicity of ChAdOx1 resulting in participant IgG levels waning below protective thresholds faster than in BNT162b2 recipients^13^.

In summary, the data generated here add detail to the evidence of broader immune responses to infection and vaccination beyond the comparatively well-characterised anti-S IgG and neutralising antibody levels as markers of protection against SARS-CoV-2 BIs. Additionally, our findings support the notion that it is not only the magnitude of T-cell responses generated by hybrid immunity that improves protection against BI, but also their breadth and quality. This reinforces the need for next generation vaccines that elicit hybrid-mimicking immune responses for more robust and sustained protection. Better understanding the immune markers correlated with protection will also be important for the rapid validation of these new vaccines at a point in the pandemic when placebo-controlled vaccine trials are no longer practical.

## Data Availability

All data produced in the present study are available upon reasonable request to the authors.

## ACKNOWLEDGEMENTS

We are extremely grateful to all the families who took part in this study, the midwives for their help in recruiting them, and the whole ALSPAC team, which includes interviewers, computer and laboratory technicians, clerical workers, research scientists, volunteers, managers, receptionists and nurses.

We wish to acknowledge the assistance of Dr Andrew Herman, Helen Rice, and the University of Bristol Faculty of Health and Life Sciences Flow Cytometry Facility. This work was undertaken with support of the Elizabeth Blackwell Institute (EBI) Mechanisms to Populations Research Strand that promotes interdisciplinary research between fundamental bioscience and population health. The support from the EBI and the University’s Alumni and Friends funded the purchase of the CTL ELISpot reader and ELISA microplate washer, and ELISpot peptide pools used in this study. Thank you to the Bristol UNCOVER group for helpful discussions on assay development and the interpretation of results.

## AUTHOR CONTRIBUTIONS

Conceptualisation, AF NJT LW AH LR; Methodology, HEB MS OF EM EO REM AT AH; Validation, HEB MS OF EM EO AT AF; Formal Analysis, HEB MS OF REM MK AH; Investigation, HEB MS OF EM KE EO BH DD AT BM; Resources, KG NDB PK AB IB AMT; Data Curation, KN FR; Writing – Original Draft, HEB MS LW AH LR; Writing – Review and Editing, OF EO REM PK AG AF NJT; Visualisation, HEB MS OF; Supervision, AMT IB AD AG AF LW AH LR; Project Administration, JO SR LM LW AH LR; Funding Acquisition, AF SR LM NJT LW AH LR

## METHODS

### ALSPAC

The Avon Longitudinal Study of Parents and Children (ALSPAC) is a birth cohort study ^73–76^. Pregnant women resident in Avon, UK with expected dates of delivery between 1st April 1991 and 31st December 1992 were invited to take part. A total of 14,541 pregnancies were initially enrolled, with 13,988 children who were alive at 1 year of age. ALSPAC now comprises three generations: the original pregnant women with the biological fathers and other carers/partners (G0), the cohort of index children (G1), and the offspring of the index children (G2). This has generated a wealth of biological, genetic and phenotypic data spanning the lifetime of these individuals. Including additional participants recruited in the interim period, the total sample size for analyses using any data collected after the age of seven is 15,447 pregnancies, resulting in 15,658 foetuses. Of these 14,901 children were alive at 1 year of age. 12,113 G0 partners have been in contact with the study, of which 3,807 are currently enrolled. Please note that the study website contains details of all the data that are available through a fully searchable data dictionary and variable search tool ^77^. From the beginning of the COVID-19 pandemic, ALSPAC sought to utilise their unique expertise and infrastructure to contribute to SARS-CoV-2 research efforts through collection of biological samples and questionnaire data from their cohort of well characterised participants.

### Study design

In October 2020, ALSPAC undertook 5200 serological SARS-CoV-2 spike-specific lateral flow tests (Fortress Diagnostics, Antrim Northern Ireland) on G0 and G1 cohort participants ^78^. Participants with evidence of a previous SARS-CoV-2 infection based on a positive IgG result on this LFT, and/or a positive SARS-CoV-2 PCR result from linked UK Health Security Agency (UKHSA) data, were invited to take part in the study (n=124). Two control groups with negative serological results, and no documented positive PCR test, were recruited alongside these participants. The first control group (n=93) were also age, sex, and symptom (anosmia) matched to those with a history of SARS-CoV-2 infection. Participants in the second control group (n=103) were selected on the basis of not having reported anosmia. Full details of the recruitment methodology, alongside a detailed characterisation of the cohort at clinic 1 (pre-vaccination), are described in Mitchell *et al* ^30^. In brief, participants attended up to 3 clinics in December 2020 (clinic 1), March 2021 (clinic 2) and June 2021 (clinic 3) where they provided venous blood and saliva samples. Additional participants were recruited at clinics 2 and 3 to maintain numbers and account for those who withdrew from the study. Health and lifestyle information was gathered through online questionnaires. For the purposes of the analyses presented in this study, the two control groups have been combined as no significant differences in baseline antibody or T-cell measures were detected between the two original groups ^30^.

### Ethics

Ethical approval for the study was obtained from the ALSPAC Ethics and Law Committee and the Local Research Ethics Committees (NHS REC 20/HRA/4854). Consent for biological samples was collected in accordance with the Human Tissue Act (2004). Informed consent for the use of data collected via questionnaires and clinics was obtained from participants following the recommendations of the ALSPAC Ethics and Law Committee at the time.

At age 18, study children were sent ‘fair processing’ materials describing ALSPAC’s intended use of their health and administrative records and were given clear means to consent or object via a written form. Data were not extracted for participants who objected, or who were not sent fair processing materials. Ethical approval was obtained from the ALSPAC Law and Ethics committee and local research ethics committees (NHS Haydock REC 10/H1010/70).

### Sample collection and processing

Peripheral blood mononuclear cells (PBMCs) were obtained from up to 3 x 10 ml EDTA tubes per participant using standard density gradient separation techniques. Briefly, samples were kept at room temperature for up to 3 hours after drawing blood. Blood was diluted 1:1 with Phosphate Buffered Saline (PBS) containing 1% Fetal calf serum (FCS). Diluted blood was separated using a Ficoll gradient, centrifuging at 1000g for 10 mins at room temperature. PBMCs were washed in PBS/1% FCS, centrifuging at 700g for 10 mins at room temperature, and washed again with PBS/1% FCS, centrifuging at 400g for 10 mins at room temperature. Cell pellets were resuspended in freezing mix (90% FCS/10% DMSO) at a concentration of 13-15×10^6^ cells per mL. Cells were frozen overnight in an alcohol bath to control freezing rate. PBMCs were then transferred to liquid nitrogen cryotanks for long-term storage. Serum tubes were left to clot, centrifuged at 1,500g for 10 mins at 18-25°C, then serum was removed, aliquoted, and stored at -80 °C.

Participants provided neat saliva directly into a sterile collection tube. Particulate matter was removed by centrifugation at 13,000g for 10 minutes. Samples were heat-inactivated at 56°C for 30 minutes prior to ELISA analysis.

### ELISAs

SARS-CoV-2-specific anti-Spike (S) IgA and IgG in serum and saliva were measured by ELISA. Samples were run in duplicate at a single optimised dilution and reported in BAU/mL following calibration of an internal standard to the WHO/NIBSC reference control. Serum anti-nucleocapsid (N) and anti-S pan Ig were utilised as screening ELISAs to report a positive/negative result for previous SARS-CoV-2 infection with data presented as normalised optical density (OD) measurement relative to an internal control. Thresholds for positivity were calculated on a large sample of PCR-confirmed and pre-pandemic samples. Full details of these methods are published for serum ^79^, and saliva ^80^, respectively.

### Synthetic peptides

Peptides used for PBMC stimulations in the ELISpot assays are listed in Supplementary Table S1. 15-mer peptides overlapping by 10 amino acids and spanning the sequences of the SARS-CoV-2 S, N, Membrane (M) and Envelope (E) protein were purchased from Mimotopes (Australia). The purity of the peptides was >80%. 15-18mer peptides overlapping by 10 amino acids and spanning sequences of SARS-CoV-2 NSP1-16, ORF3, ORF6, ORF7 and ORF8 were also purchased). The purity of the peptides was ∼70%. The combination of peptides and identity of the peptide pools are described in Methods Table 1. For PBMC stimulation in ELISpot assays, pools of a maximum of 127 peptides were added per well as follows: Spike was divided into 2 pools (S1; 126 peptides and S2; 127 peptides); for smaller peptide regions, peptides were combined in the same peptide pools as follows: NSP1 and NSP2 (NSP1+2), NSP5 and NSP6 (NSP5+6), NSP15 and NSP16 (NSP15+16). All other peptides were pooled into one mixture and tested in individual ELISpot wells.

### Enzyme-linked immune absorbent spot assay (ELISpot)

Human IFN-γ ELISpot assays were performed using a Human IFN-γ ELISpot BASIC kit (Mabtech). MSIP4W10 PVDF plates (Millipore) were coated with capture antibody (mAb-1-D1K; 15 μg/mL) and incubated overnight at 4 °C in carbonate bicarbonate buffer (Sigma Aldrich). Cryopreserved PBMC were thawed then rested at 37 °C/5% CO_2_ for 5-6 hours. Coated plates were washed 5 times in sterile PBS and blocked for 1-2 hours using R10 medium (0.2 µm filtered RPMI 1640 medium supplemented with 10 % FBS, 2 mM glutamine, penicillin ((100 units/ml) and streptomycin (100 μg/ml)). 4 x 10^5^ PBMCs were added to each well in the plate, with or without peptide pools (as indicated) in a total assay volume of 100 µl in R10. PBMC incubated with R10 medium alone were used as negative (unstimulated) controls and were performed in duplicate. Peptide pools spanning S1, S2, M, N, E, ORF1 (NSP1+2, NSP3A, NSP3B, NSP3C, NSP4, NSP5+6, NSP7-11, NSP12A, NSP12B, NSP13, NSP14, NSP15+16), ORF3, ORF6, ORF7 and ORF8 were used at a final concentration of 2 µg/ml. PBMCs from cases were tested against all the above peptide pools, while PBMCs from controls were tested against the following peptide pools only: S1, S2, M, N, NSP3B, NSP12A, NSP12B, NSP7-11, NSP13 and NSP15+16. PBMC stimulated with anti-CD3 antibody (Mabtech, Mab CD3-2; final concentration 0.1% v/v) were used as a positive control for each participant, with 1-4 x 10^5^ PBMCs used per well. Positive control and peptide stimulated wells were performed in singlet. Plates were incubated for 16-18 hours at 37°C/5% CO_2_ then developed as per manufacturer’s instructions. Developed plates were protected from light and air-dried for 48 hours before image acquisition using a CTL ImmunoSpot S6 Ultra-V Analyzer. Spot forming units (SFU) were calculated using the ImmunoSpot S6 Ultra-V Analyzer Basic Count function after image acquisition using optimised counting parameters that were applied across all participants. Spot counts were enumerated for each peptide pool by subtraction of average background (calculated from duplicate unstimulated wells). Counts were expressed as SFU per million (10^6^) PBMC after multiplication by 2.5 following background subtraction. Negative values after background subtraction were adjusted to zero ^29^. Participants were excluded if the spot count in unstimulated wells exceeded 95 SFU per million PBMC or if no spots were observed in the positive control wells. Where spot formation was too dense to accurately enumerate using standardised counting parameters (TNTC; too numerous to count), affected wells were excluded unless contemporaneous assessment of IFN-γ production by flow cytometry confirmed antigen specific response. In these cases, TNTC values were given the raw value equivalent to the largest spot count accurately counted for a peptide pool (320 SFU per well).

### Intracellular Cytokine Staining (ICS)

PBMCs were thawed and rested overnight in AIMV 2% FCS then incubated with or without peptide pools from SARS-CoV-2 S1/S2, M, N, NSP3B (all 1 μg/ml), or with PMA/ionomycin (PMA 10 ng/ml, ionomycin 100 ng/ml, Sigma-Aldrich) for 5 hr at 37°C in the presence of brefeldin A (BD, 5 μg/ml). To assess degranulation, anti CD107a-FITC antibody was added to the cells at the beginning of the stimulation. Cells were stained with a viability dye Zombie Aqua (BioLegend) for 10 min at room temperature and then with antibodies targeting surface markers (20 min 4°C, diluted in PBS 1% BSA; Sigma-Aldrich). Cells were fixed overnight in eBioscience Foxp3/Transcription factor fixation/permeabilization buffer (Invitrogen), and intracellular staining was performed for detection intracellular cytokines, including IFN-γ, TNF-α, IL-2 and MIP1β (30 min 4°C). Four samples were excluded from further CD4^+^ T-cell analysis due technical issues. Data were acquired on a BD LSR Fortessa X20 and analysed using FlowJo software v10.8.1. Results were obtained after subtraction of the values in the corresponding unstimulated well. A complete list of antibodies is included in Methods Table 2. ICS was performed on samples from 69 individuals in the case group, selected based on having a detectable T-cell response by IFN-γ ELISpot at baseline (i.e. clinic one) for at least one of the SARS-CoV-2 peptide pools tested and for whom we had additional cryopreserved PBMC vials available at baseline.

### Pseudoneutralisation

Serum neutralisation was expected to positively correlate with levels of anti-Spike antibody binding results. Accordingly, all samples from participants in the case group corresponding to anti-Spike pan-Ig ELISA results above a normalised threshold of 0.5 were included. For pseudovirus assays, Wuhan-Spike-harbouring pseudovirus (luciferase-expressing vesicular stomatitis virus, VSV-S-FLuc) was generated and used to assess serum antibody neutralisation of VSV-S-FLuc entry into Vero ACE2 TMPRSS2 (VAT) cells as described previously ^79^. Briefly, serum dilutions starting at 1/40 followed by eight 2.5-fold titrations were plated in triplicate in 96-well plates, alongside three wells each of 1/25 dilutions of known neutralising and non-neutralising controls (corresponding to 16,000 RLU of luminescence when mixed with VSV-S-FLuc). WT Wuhan spike pseudotyped VSV was added to each well and incubated for an hour. Well-mixtures were added to black, microscopy 96-well plates, seeded with 10,000 Vero ACE2 TMPRSS2 (VAT) cells per well. Luminescence measurements were taken 16 hours after infection, using the ONE-Glo Luciferase Assay System.

### Data analysis

Study data were collected and managed using REDCap (Research Electronic Data Capture) electronic data capture tools hosted at the University of Bristol. REDCap is a secure, web-based software platform designed to support data capture for research studies ^81^.

Participant data was excluded from the analysis if it was subsequently established that a participant was vaccinated as part of an unlicensed COVID-19 vaccine trial prior to enrolment in this study. In order to focus on the immune responses to COVID-19 vaccines, data was also removed if no corresponding information provided on the COVID-19 vaccination status of the participant at a particular clinic visit. Where a participant provided samples at more than one clinic after a specified number of vaccine doses (including pre-vaccination), only the data from the earliest sampling clinic was included in the analysis.

Statistical analyses were performed using R Studio (v4.3.0), and GraphPad Prism (version 10.01). Unpaired comparisons across multiple groups were done with the Kruskal-Wallis test with Dunn’s post-test for multiple comparisons. Pairwise correlations were assessed with Spearman’s rank-order correlation (r_s_). Correlation coefficients were interpreted as: weak (r_s_=0.20-0.39), moderate (r_s_=0.40-0·59), strong (r_s_=0.60-0.79), or very strong (r_s_=0.80-1.00). The following adjusted P value thresholds were used for data visualisation: P≤0.05 (*), P≤0.01 (**), P≤0.001 (***), P≤0.0001 (****). To facilitate the presentation of data which included zero counts on log scales, zero counts were plotted as 1 (or the minimum y axis baseline, if lower) for visualisation purposes only – all statistical analyses were performed on the raw data values.

---

**Table S1:** Participant demographics. To protect participant anonymity, specific values for counts fewer than 5 are not provided. Please note that fields marked as < 5 may include zero.

|  | Cases (N=199) | Controls (N=176) |
| --- | --- | --- |
| **Sex** |  |  |
| Female / male (n) | 142 / 57 | 80 / 96 |
| **Age** |  |  |
| G0 / G1 (n) | 77 / 122 | 79 / 98 |
| Mean age (years) | 39 | 42 |
| Median age (years) | 29 | 29 |
| Age unknown (n) | 22 | 0 |
| **Ethnicity** |  |  |
| Asian/Asian British (n) | < 5 | < 5 |
| Black/African/ Caribbean/ Black British (n) | < 5 | < 5 |
| Arab or other ethnic group (n) | < 5 | < 5 |
| Mixed/multiple ethnic groups (n) | < 5 | < 5 |
| White (n) | ≥ 166 | ≥ 144 |
| Ethnicity data not available (n) | 8 | 9 |

**Table S2:** Statistical comparison of monofunctional SARS-CoV-2-specific CD4^+^ and CD8^+^ T-cells in previously infected individuals. Statistics from monofunctional SARS-CoV-2-specific CD4^+^ and CD8^+^ T-cells in previously infected individuals as presented in Figure 3. Calculated using a Kruskal-Wallis test with FDR method of Benjamini and Hochberg correction for multiple comparisons.

| Cytokines | Protein | Individual P Value /<br>Monofunctional CD4<sup>+</sup> T-cells | Individual P Value /<br>Monofunctional CD8<sup>+</sup> T-cells |
| --- | --- | --- | --- |
| MIP1β vs IL2 | S1 | 0.0966 | <0.0001 |
|  | S2 | 0.1297 | <0.0001 |
|  | M | >0.9999 | <0.0001 |
|  | N | 0.0146 | <0.0001 |
|  | NSP3B | 0.0658 | <0.0001 |
| MIP1β vs CD107a | S1 | 0.0074 | <0.0001 |
|  | S2 | 0.5881 | <0.0001 |
|  | M | <0.0001 | <0.0001 |
|  | N | 0.08 | <0.0001 |
|  | NSP3B | 0.9148 | <0.0001 |
| MIP1β vs IFNγ | S1 | 0.0227 | <0.0001 |
|  | S2 | 0.0034 | <0.0001 |
|  | M | 0.0003 | <0.0001 |
|  | N | 0.0134 | <0.0001 |
|  | NSP3B | 0.208 | <0.0001 |
| MIP1β vs TNFα | S1 | <0.0001 | 0.007 |
|  | S2 | <0.0001 | 0.002 |
|  | M | <0.0001 | <0.0001 |
|  | N | <0.0001 | <0.0001 |
|  | NSP3B | <0.0001 | 0.0011 |
| IL2 vs CD107a | S1 | 0.3092 | 0.0492 |
|  | S2 | 0.0397 | 0.6678 |
|  | M | <0.0001 | 0.0015 |
|  | N | 0.4888 | 0.3673 |
|  | NSP3B | 0.0832 | 0.9519 |
| IL2 vs IFNγ | S1 | <0.0001 | 0.1972 |
|  | S2 | <0.0001 | 0.0309 |
|  | M | <0.0001 | 0.0295 |
|  | N | <0.0001 | 0.0381 |
|  | NSP3B | 0.0019 | 0.4588 |
| IL2 vs TNFα | S1 | <0.0001 | <0.0001 |
|  | S2 | <0.0001 | <0.0001 |
|  | M | <0.0001 | 0.002 |
|  | N | <0.0001 | 0.0002 |
|  | NSP3B | <0.0001 | <0.0001 |
| CD107a vs IFNγ | S1 | <0.0001 | 0.0011 |
|  | S2 | 0.0171 | 0.0837 |
|  | M | >0.9999 | 0.3141 |
|  | N | <0.0001 | 0.0029 |
|  | NSP3B | 0.1719 | 0.4961 |
| CD107a vs TNFα | S1 | <0.0001 | <0.0001 |
|  | S2 | <0.0001 | <0.0001 |
|  | M | 0.1842 | 0.9218 |
|  | N | <0.0001 | <0.0001 |
|  | NSP3B | <0.0001 | <0.0001 |
| IFNγ vs TNFα | S1 | <0.0001 | 0.005 |
|  | S2 | <0.0001 | 0.0113 |
|  | M | 0.0566 | 0.3636 |
|  | N | 0.0001 | 0.1086 |
|  | NSP3B | 0.002 | 0.0004 |

**Table S3:**
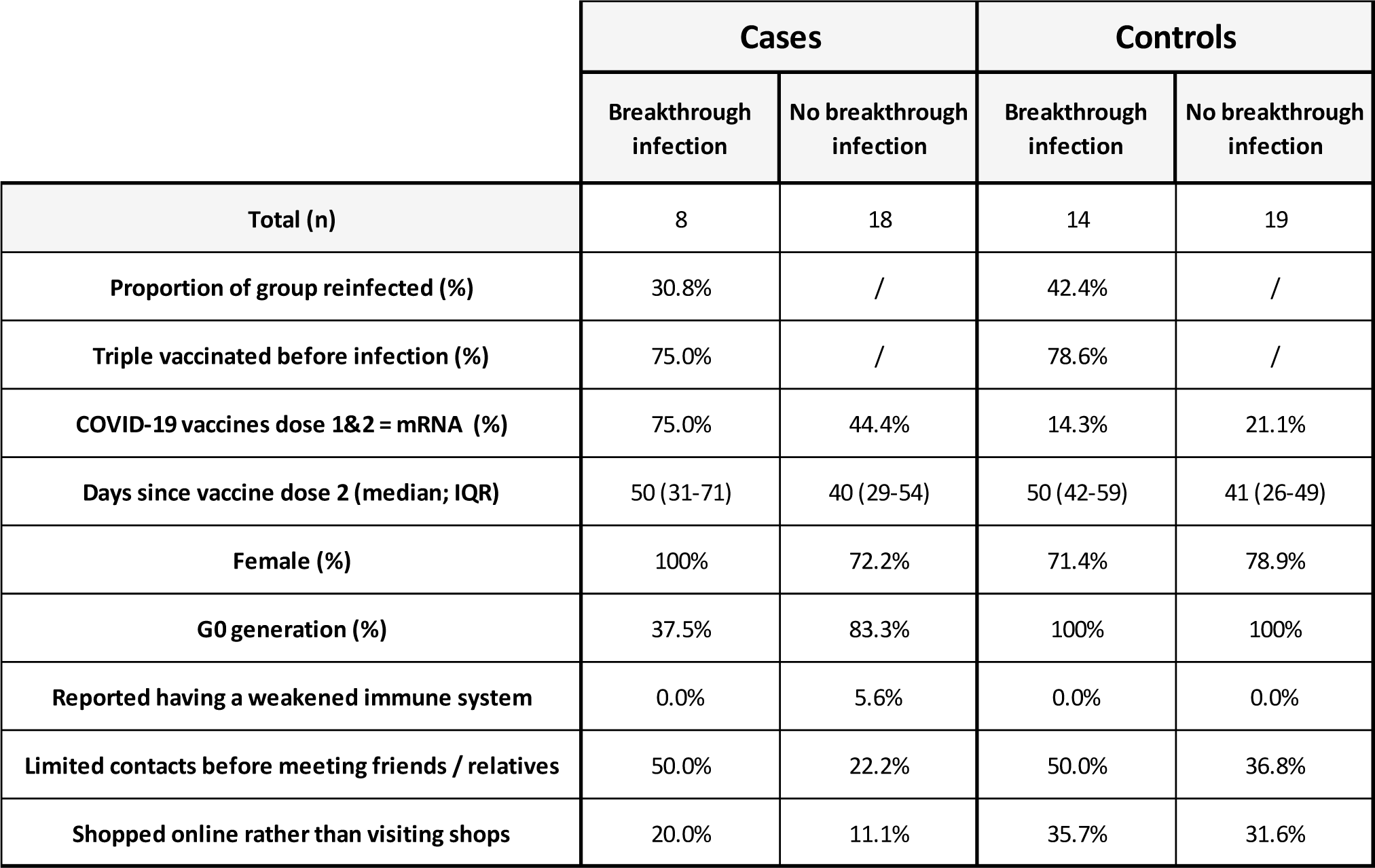
Characteristics of the infection susceptibility groups. Participants were classified as cases based on a PCR-confirmed SARS-CoV-2 infection prior to their second vaccine dose, and/or positivity on the serum anti-Spike and/or anti-Nucleocapsid protein pan Ig screening ELISAs at clinic 1 (prior to vaccination). In each of the case and controls groups, participants were stratified into those who did and didn’t self-report a SARS-CoV-2 infection in the 8 months after clinic 3 (July 2021 – March 2022). Participants self-reported information on whether they identified as having a weakened immune system / increased susceptibility to infection at the beginning of the pandemic (April 2020). Data on behavioural modifications were collected at the end of the breakthrough infection monitoring period (March 2022). Participants self-reported whether they felt they had been limiting their social contacts prior to meeting friends or relatives, and whether they had been shopping online rather than visiting stores.

**Supplementary Figure 1:**
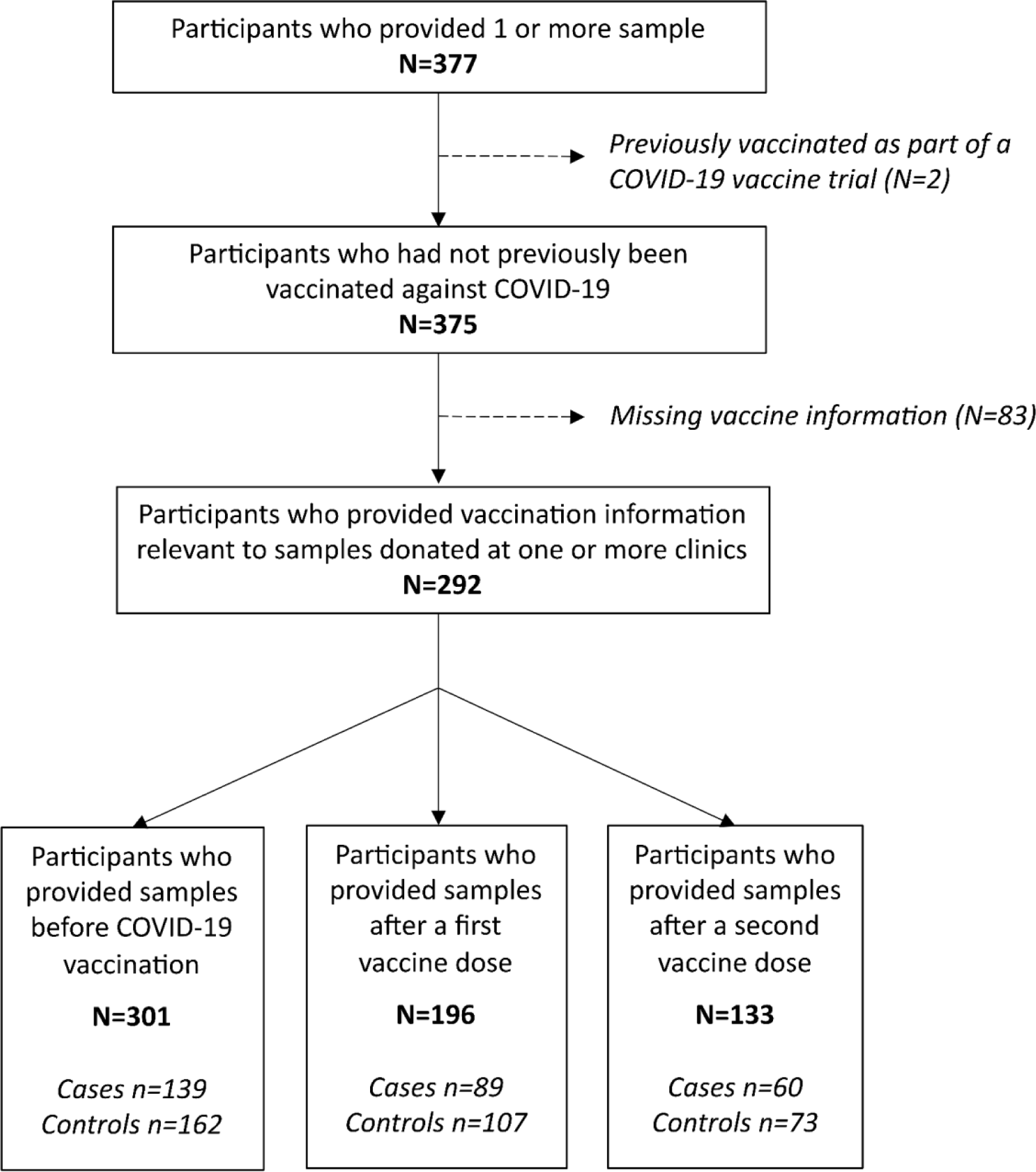
Participants inclusion flowchart. All G0 (original mothers and partners) and G1 (offspring generation) ALSPAC participants were eligible to voluntarily enrol in this study. Participant data was subsequently excluded from the analysis if they reported a previous COVID-19 vaccination. In order to focus on the immune responses to COVID-19 vaccines, data was also removed if there was no corresponding information provided on the COVID-19 vaccination status of the participant at a particular timepoint.

**Supplementary Figure 2:**
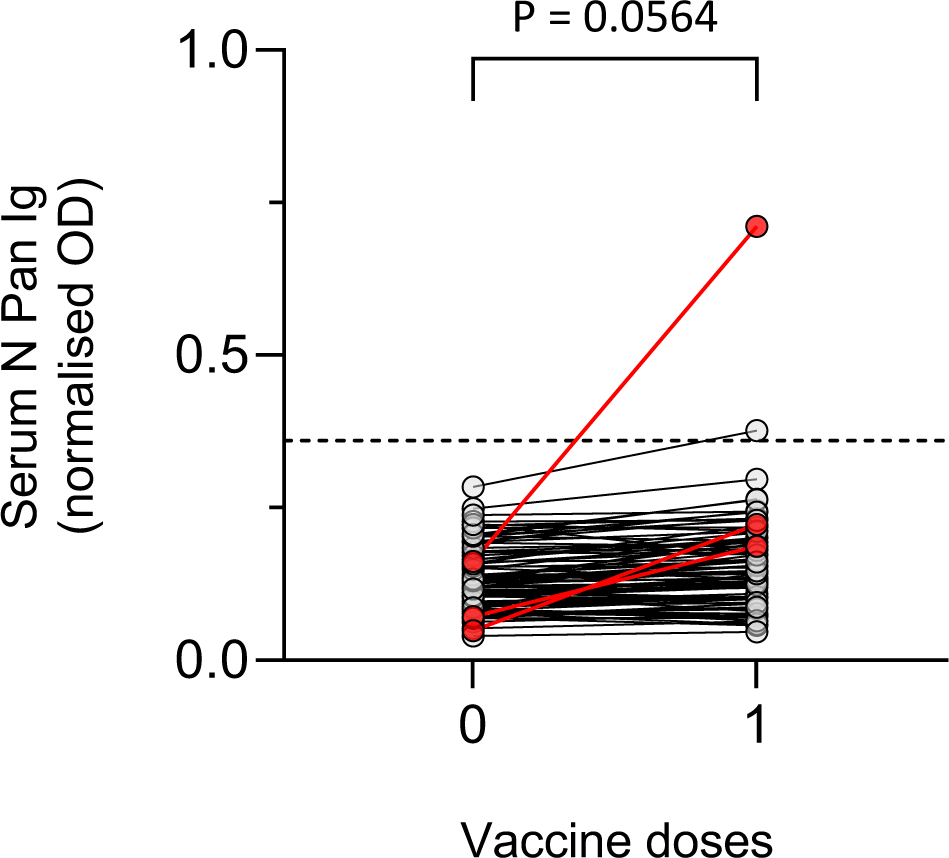
Anti-N pan Ig levels in control participants before and after vaccine dose 1. Serum anti-Nucleocapsid (N) pan Ig antibody levels in serum were measured by in-house ELISA. Results are presented as normalised OD relative to an internal control. Data shown represent all participants classified as controls with data available both before and after the first vaccine dose (n=95). Participants with a ≥ 2-fold increase in N pan Ig levels from baseline to post-dose 1 are highlighted in red (n=3). Responses before and after vaccination were compared using a Mann Whitney test.

**Supplementary Figure 3:**
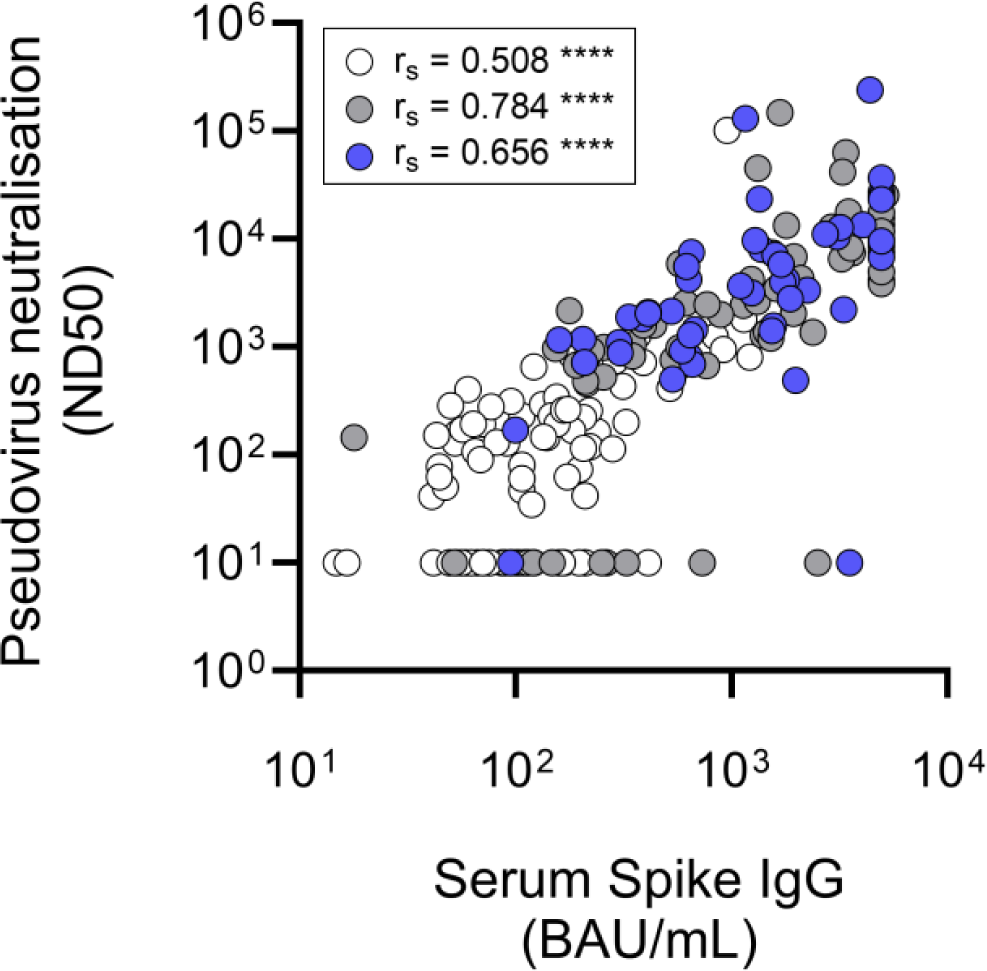
Correlation between SARS-CoV-2 Spike-specific serum IgG and pseduoneutralising antibody levels. Anti-Spike IgG levels in serum were measured by in-house ELISA and reported in BAU/mL following calibration of the assay to the WHO/NIBSC reference standard. Pseudoneutralising antibody titres against ancestral Spike were measured in serum. Spearman’s rank (r_s_) correlations were calculated at baseline (white; n=99), post dose 1 (grey; n=61) and post dose 2 (blue; n=49). P values were categorised as ≤0.05 (*), ≤0.01 (**), ≤0.001 (***), ≤0.0001 (****).

**Supplementary Figure 4:**
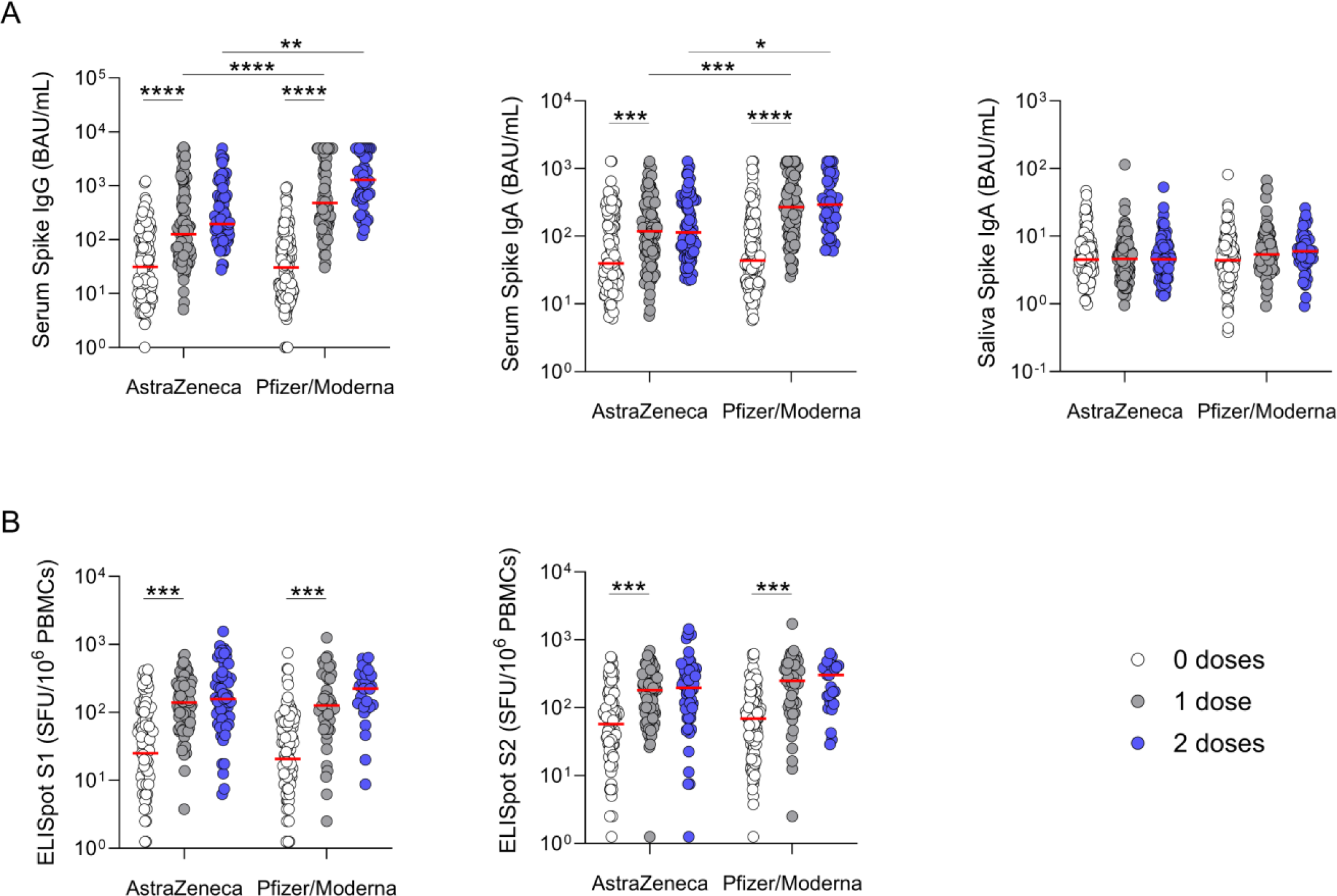
Antibody and T cell responses by COVID-19 vaccination type. Participants were stratified based upon the type of COVID vaccine received; AstraZeneca ChAdOx1 whole virion vaccine, or one of two mRNA-based vaccines (Pfizer/BioNtech BNT162b2 or Moderna mRNA-1273). All participants received homologous first and second doses. (A) Antibody levels in serum and saliva were measured by in-house ELISA. Assay thresholds were set using samples taken following recent SARS-CoV-2 infection and pre-pandemic controls to achieve 99% specificity. Results are reported in binding antibody units (BAU)/mL following calibration of the assay to the WHO/NIBSC reference standard. (B) Magnitude of SARS-CoV-2 specific T cell response against Spike (pool-1 and 2) peptide pools measured via ELISpot assay. Shown are comparison of cells producing IFN-γ after stimulation of PBMCs with overlapping peptides spanning the indicated proteins. Results are expressed as Spot Forming Units (SPU) relative to 1×10^6^ PBMCs after subtraction of average background (calculated from duplicate unstimulated wells). Negative values after background subtraction were adjusted to zero. Participants were excluded from the dataset due to high background (>95 SFU per million PBMC). White = baseline measurement; grey = post vaccine dose 1; blue = post vaccine dose 2. Red bars represent the median of each group. Unpaired comparisons were performed using Kruskal-Wallis test with Dunn’s correction for multiple comparisons. Within each of the case and control groups, responses were compared between 0 and 1, and 1 and 2 vaccine doses. Responses between groups were compared after each dose. Statistics are only displayed for comparisons where P≤0.05. P values were categorised as ≤0.05 (*), ≤0.01 (**), ≤0.001 (***), ≤0.0001 (****).

**Supplementary Figure 5:**
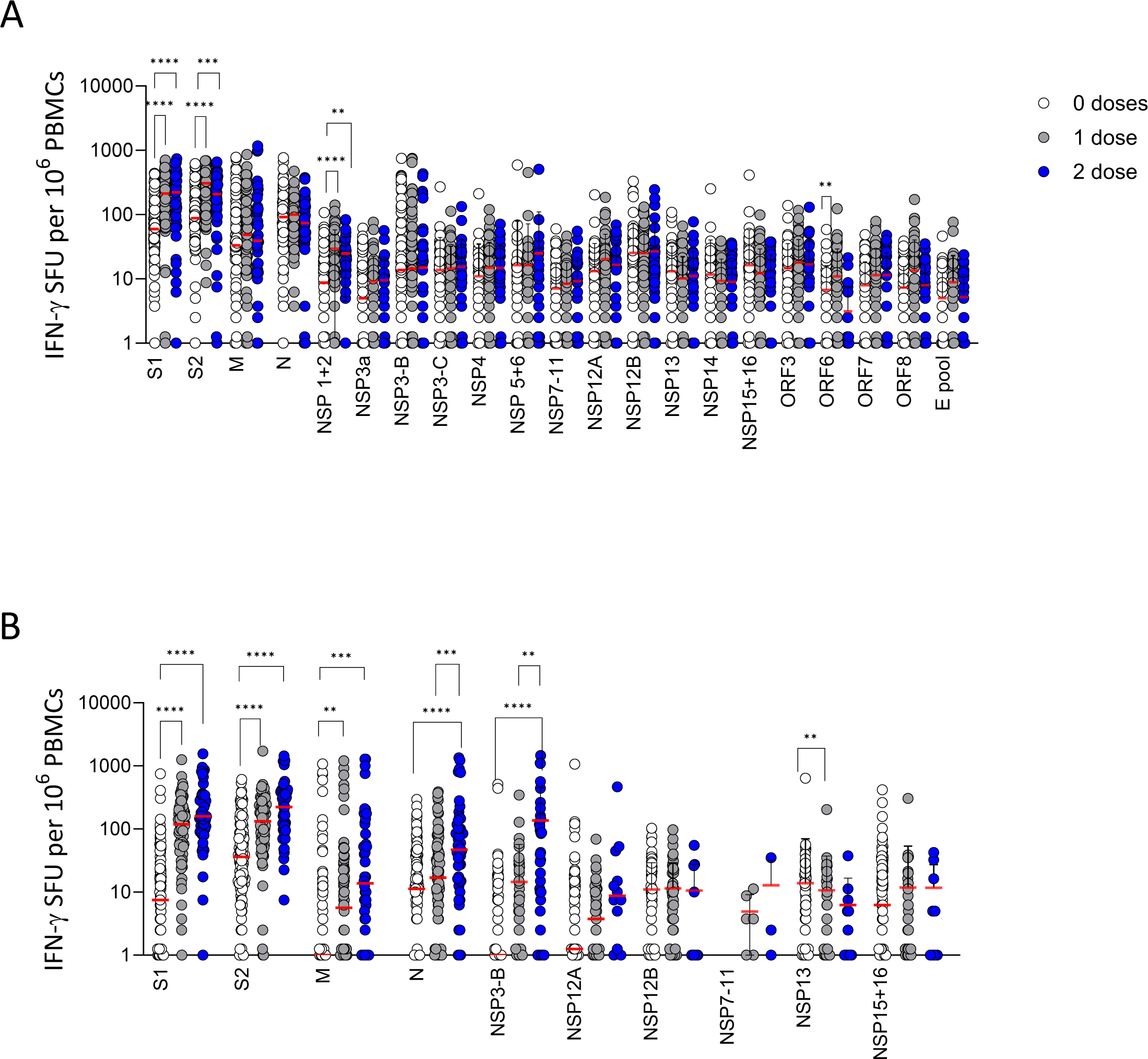
Magnitude of T-cell response following COVID-19 vaccination. Ex-vivo IFN-γ ELISpots showing the overall magnitude and breadth of effector T-cell responses to SARS-CoV-2 proteins following vaccination. (A) Responses in cases (n=108), and (B) responses in controls (n=132). Results are expressed as Spot Forming Units (SFU) per 1×10^6^ PBMCs after subtraction of average background (calculated from duplicate unstimulated wells). Red bars represent median T-cell responses. Structural proteins: Spike (S1 and S2), M, N and E, non-structural: NSP1-2, NSP3A, NSP3B, NSP3C, NSP4, NSP5-6, NSP7-11, NSP12A, NSP12B, NSP13, NSP14 and NSP15-16, and accessory proteins: ORF3, ORF6, ORF7 and ORF8. Significance was determined using a Kruskal-Wallis test with Dunn’s correction for multiple comparisons; where P ≤0.05 (*), ≤0.01 (**), ≤0.001 (***), ≤0.0001 (****).

**Supplementary Figure 6:**
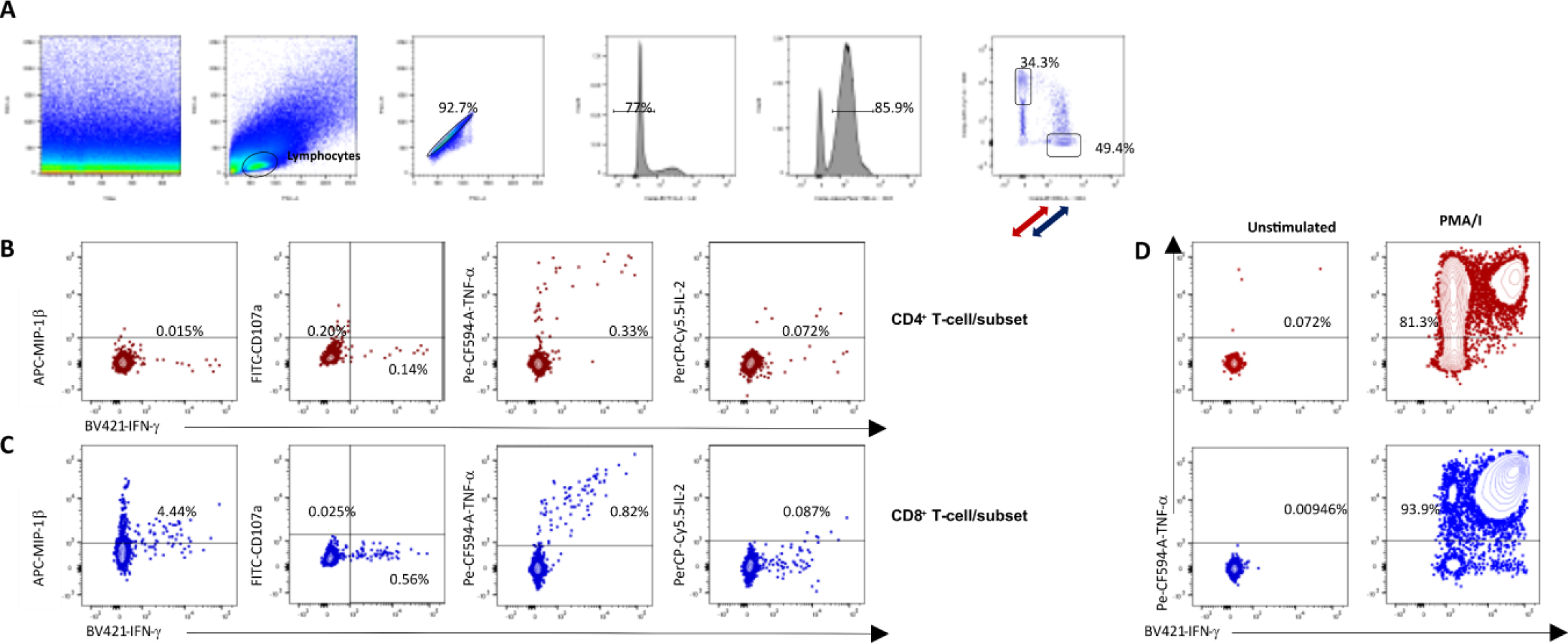
Gating-strategy for the identification of SARS-CoV-2-reactive cytokine producing CD4^+^ and CD8^+^ T-cells. PBMCs were stimulated with SARS-CoV-2-overlapping-peptide-pools (Spike, M, N and NSP3B) for 6h after overnight resting. The gating strategy for CD4^+^ and CD8^+^ T-cells is shown panel A. Dot plots are representative of cytokine secretion by CD4^+^ T-cells (B) and CD8^+^ T-cells (C) specific for the Membrane (M) protein. These cells were further analysed for the expression of interleukin (IL) 2, interferon γ (IFN-γ), tumour necrosis factor (TNFα), MIP1β and degranulation (CD107a) (panel B/C). The unstained control and PMA/I stimulated sample were used as a negative and positive control for ICS assay, respectively (D).

**Supplementary Figure 7:**
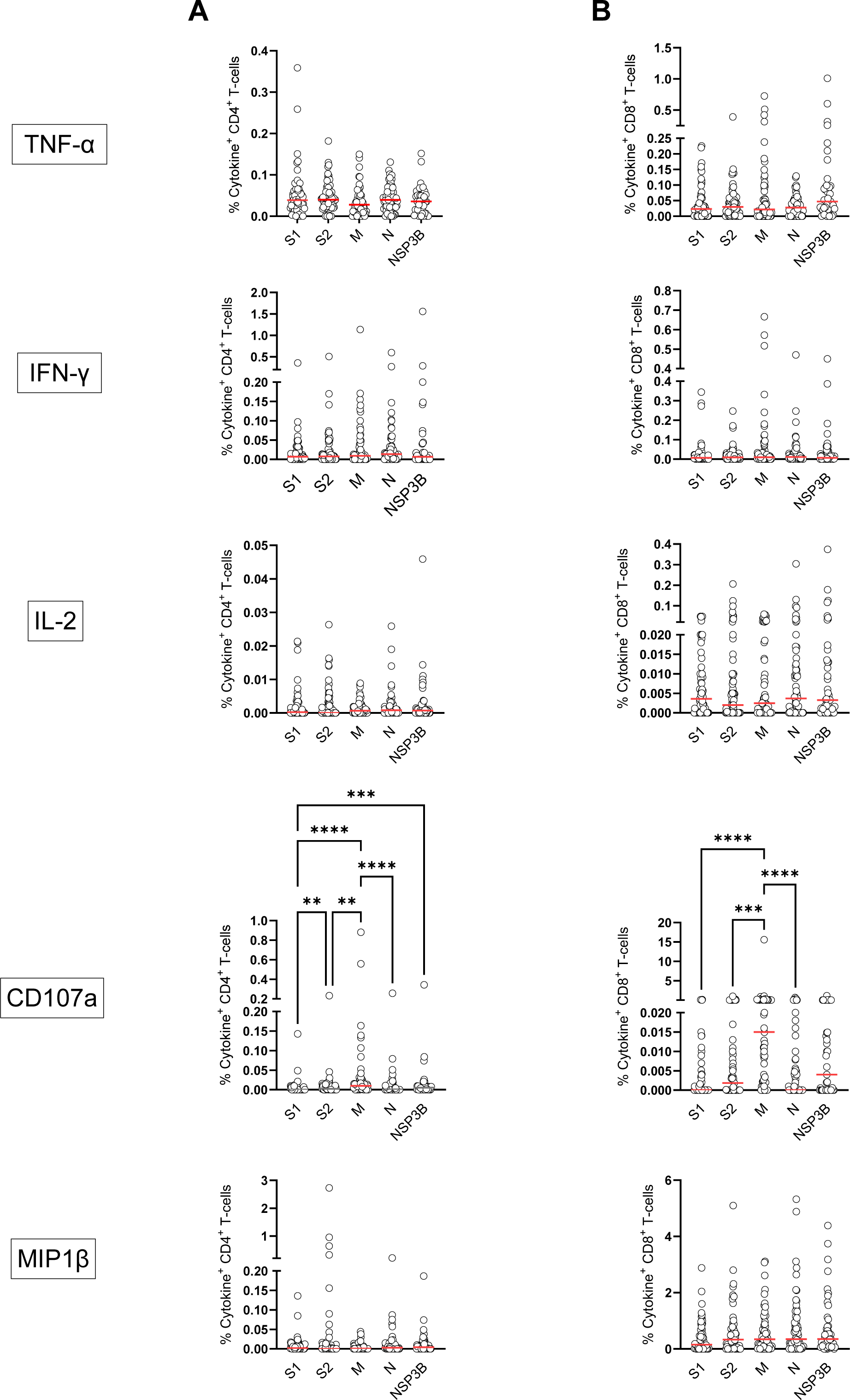
Frequency of monofunctional SARS-CoV-2-specific CD4^+^ and CD8^+^ T-cells in previously infected individuals. Percentage of cytokine-producing T-cells was measured by intracellular cytokine staining and flow cytometry. Shown are the percentages of monofunctional CD4^+^ (A) and CD8^+^ (B) T-cells producing IFN-γ, TNFα IL-2, MIP1β or CD107a (single positive for each) after stimulation with the indicated SARS-CoV-2 peptide pools. Statistics were calculated using a Kruskal-Wallis test with Dunn’s correction for multiple comparisons; where P ≤0.05 (*), ≤0.01 (**), ≤0.001 (***), ≤0.0001 (****).

**Supplementary Figure 8:**
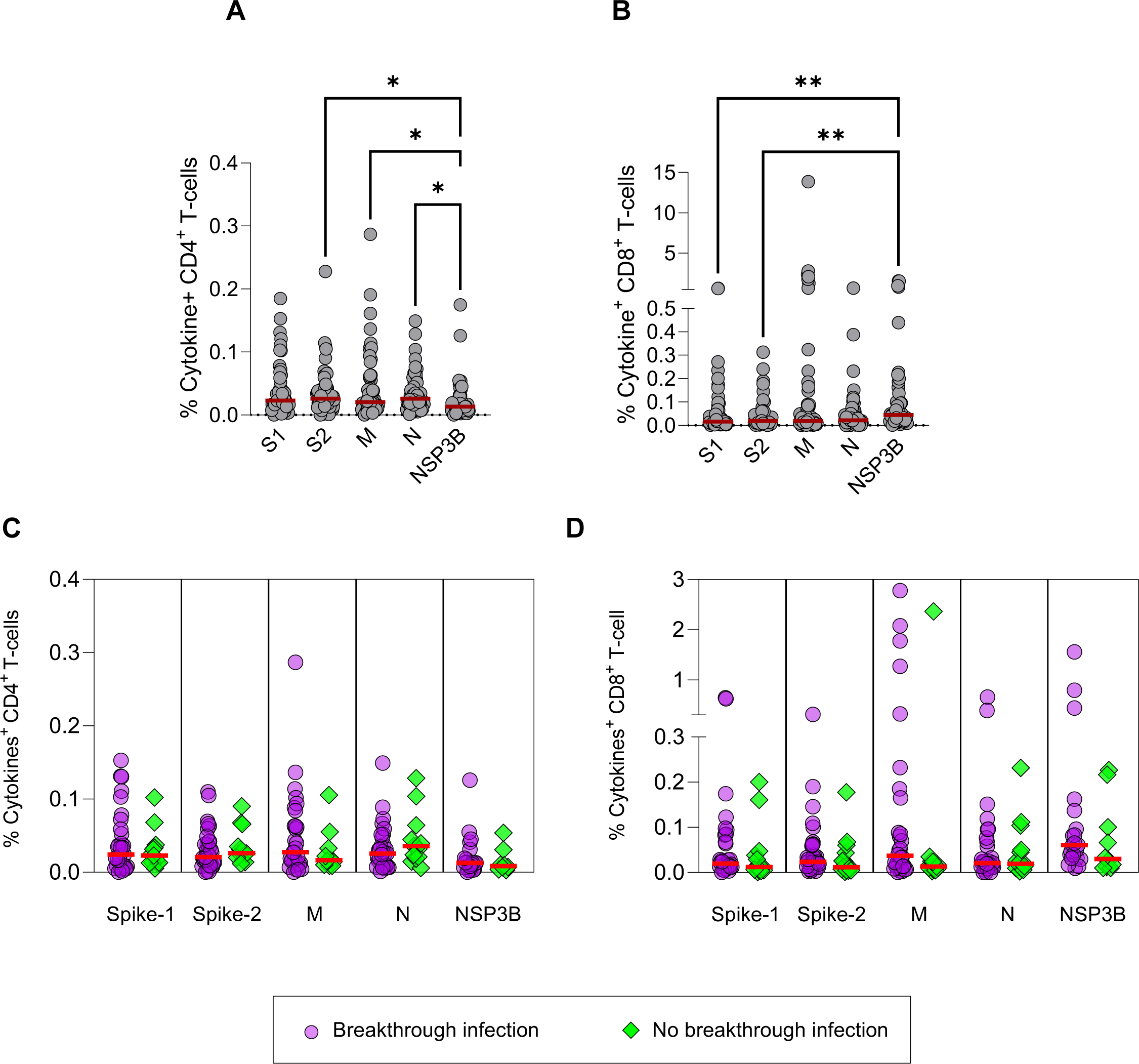
Frequency of polyfunctional SARS-CoV-2-specific CD4^+^ and CD8^+^ T-cells in previously infected individuals. (A-B) Frequency of polyfunctional SARS-CoV-2-specific CD4^+^ (left panel) and CD8^+^ (right panel) T-cells targeting peptide pools from Spike and non-Spike proteins (M, N and NSP3B). Statistics were determined using a Kruskal-Wallis test with Dunn’s correction for multiple comparisons. (C-D) Frequency of polyfunctional SARS-CoV-2-specific CD4^+^ (left panel) and CD8^+^ (right panel) T-cells targeting SARS-CoV-2 peptide pools in individuals who experienced BI infection versus those who did not. Red bars represent median responses. Statistics were calculated by t-test (Mann-Whitney test).

**Supplementary Figure 9:**
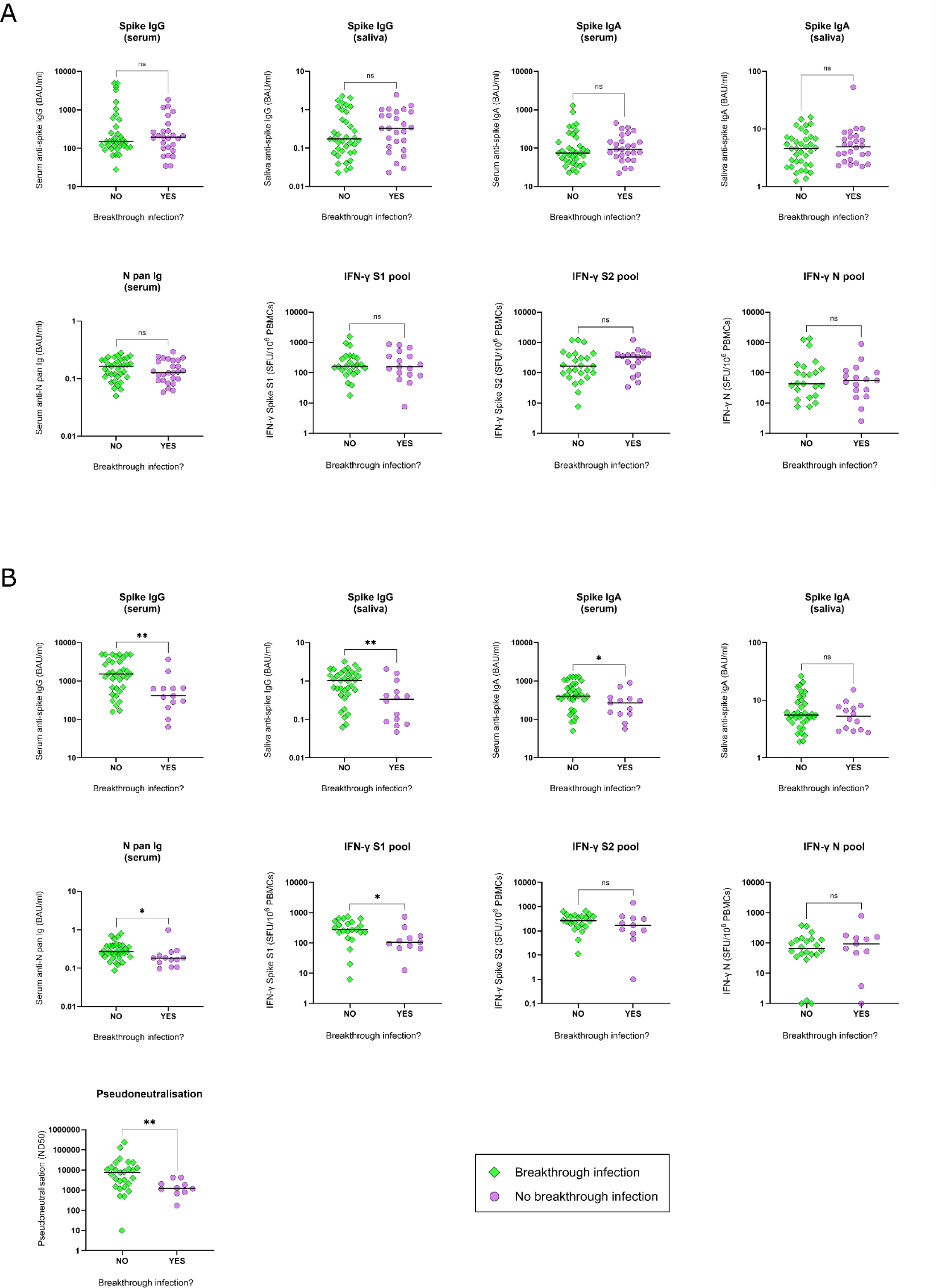
Comparison of post-vaccination dose 2 antibody and T-cell levels in participants with and without a reported SARS-CoV-2 breakthrough infection. Post-second vaccination immune responses in participants who self-reported a SARS-CoV-2 infection in the subsequent 8-months (breakthrough infection; BI) and those who did not (no breakthrough infection). (A) SARS-CoV-2-naïve individuals with (n=14) or without (n=19) a BI. (B) Previously SARS-CoV-2-infected individuals with (n=8) or without (n=18) a BI. Bars represent the median response. Groups were compared using a Mann-Whitney test, with statistics categorised as: P >0.05 (ns), P ≤0.05 (*), ≤0.01 (**), ≤0.001 (***), ≤0.0001 (****).

**Supplementary Figure 10:**
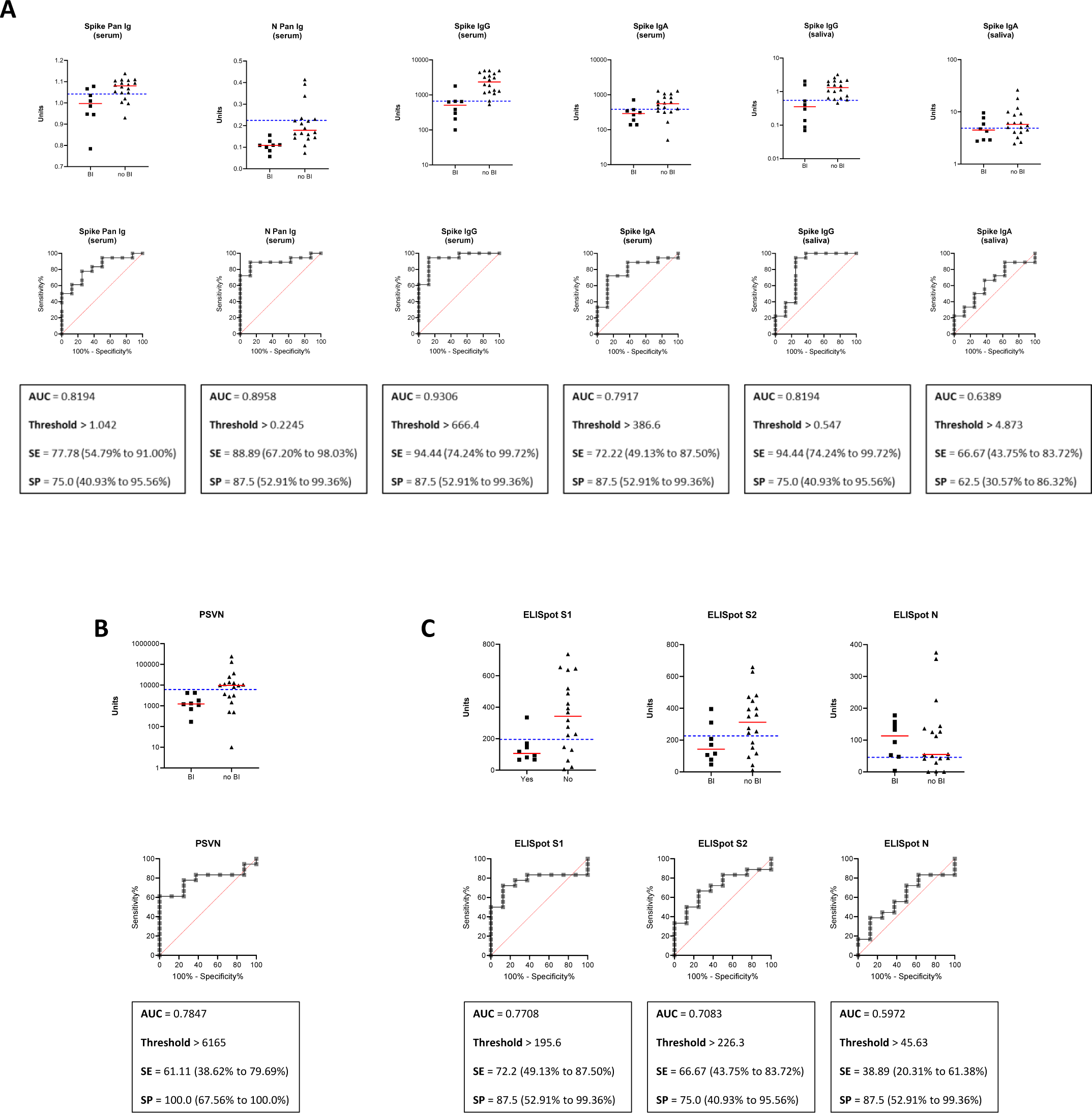
Breakthrough infection susceptibility threshold setting. Participants with a history of SARS-CoV-2 infection were stratified into two groups based upon whether they went on to be re-infected in the 8-month period after the study end date (had a breakthrough infection: yes /no). Receiver Operator Characteristic (ROC) curves were plotted for serum and saliva binding antibody measures (A), serum pseudoneutralising antibody (B), and IFN-γ producing T-cells (C). Youden’s index thresholds were then calculated for each individual measure and are represented by the blue dotted lines on each graph. The sensitivity and specificity parameters for each threshold are presented alongside 95% confidence intervals calculated using the Wilson/Brown method. AUC; area under curve, SE; sensitivity, SP; specificity. Yes BI (n=8), no BI (n=18).

**Supplementary Figure 11:**
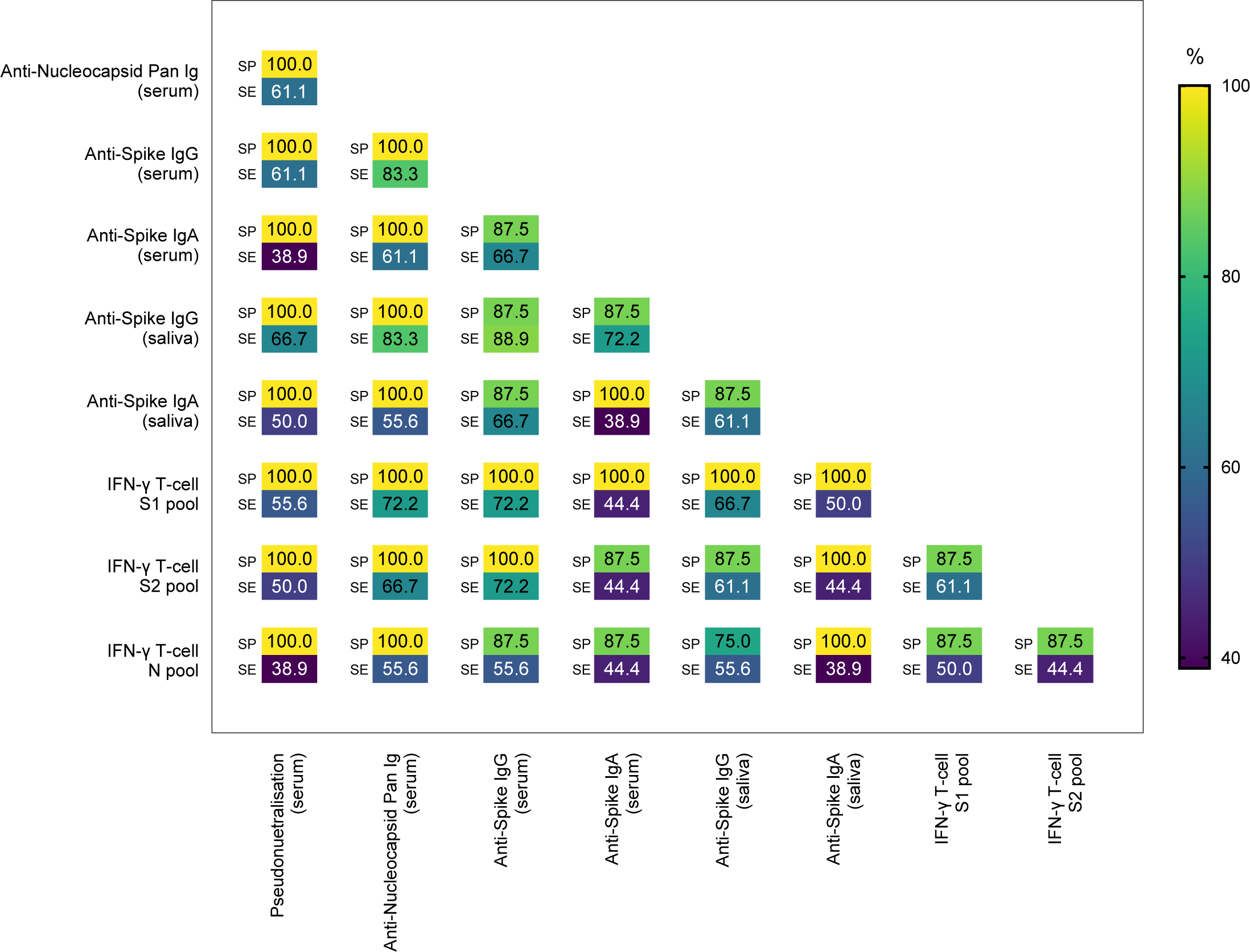
Performance of combined thresholds for breakthrough infection susceptibility. Participants with a history of SARS-CoV-2 infection were stratified into two groups based upon whether they went on to be re-infected in the 8-month period after the study end date (had a breakthrough infection, BI). Thresholds antibody and T-cell levels that were associated with protection against breakthrough infection were calculated using the Youden’s index method (figure S10 for full derivation). Values represent the specificity (SP) and sensitivity (SE) estimates for combinations of two individual thresholds.

